# Machine Learning–Driven Drug Optimization for Typhoid Fever Based on Patient Profiles

**DOI:** 10.64898/2025.12.03.25341570

**Authors:** Ssemuyiga Charles, Elminah Saru, Yusuf Abbas Aleshinloye

**Affiliations:** PharmaQsar Bioinformatics Firm, Kampala, Uganda; Department of Public Health, School of Public Health, Kampala International University, Kampala, Uganda; Department of Biochemistry and Biotechnology, Pwani University, Kenya; School of Mathematics and Computing, Kampala International University, Kampala, Uganda

**Keywords:** Typhoid fever, Machine learning, Predictive modelling, Precision medicine, and Disease Triage.

## Abstract

**Introduction:** Typhoid fever remains a major Global public health concern, with treatment outcomes dependent on antimicrobial resistance (AMR) and patient variability. Clinically determining the best medication for a certain patient can be difficult. Machine learning–based clinical decision support systems (CDSS) offer a promising avenue for improving diagnostic accuracy and guiding antibiotic selection using routinely collected clinical data

**Methods:** We developed a multi-task predictive framework using XGBoost models to predict (i) treatment outcome/success (binary), (ii) treatment duration (days), (iii) a resistance-proxy score, and (iv) suspected typhoid using clinical data; we assessed discrimination (AUC), calibration (Brier), and R² where appropriate, and used SHAP for explainability to interpret feature importance, derive patient clusters, and generate individualized model explanations. We further implemented a counterfactual drug-simulation experiment to compare actual prescribed antibiotics with predicted best options for each patient

**Results:** The treatment outcome and suspected typhoid classification models achieved an AUROC of 0.9998 ± 0.0001 and 0.9834 ± 0.0007 respectively, accurately distinguishing between successful and failed therapies and typhoid and non-typhoid cases respectively. The treatment-duration and resistance-proxy regression models demonstrated strong explanatory capacity, accounting for 80% (R² = 0.8027 ± 0.0054) and 90% (R² = 0.9014 ± 0.0036) of the variance in therapy length and inferred resistance likelihood, respectively. SHAP analysis consistently highlighted the severity group, biochemical markers as dominant predictors across models and revealed biologically coherent patient subgroups. Counterfactual drug simulations suggested that alternative antibiotics could yield higher predicted success probabilities for a subset of patients, indicating the potential value of model-informed treatment ranking

**Conclusion:** This study demonstrates the feasibility of machine learning for simulating drug choice in typhoid treatment using patient clinical profiles.

## 1.0 Introduction

Enteric fever, a systemic bacterial disease caused by *Salmonella enterica* serovars Typhi and Paratyphi (A, B, and C) collectively referred to as typhoidal Salmonella, remains a significant global health concern (1). Although the bulk of morbidity and mortality arises from *Salmonella enterica serovar Typhi /S. Typhi* (typhoid fever), infections due to *S. Paratyphi* serovars, which together account for approximately one-fifth of enteric fever cases, are emerging as a notable contributor, especially in regions transitioning to typhoid conjugate vaccine programs (1,2). In this study, we focus on typhoid fever caused by *S. Typhi*, a rod-shaped Gram-negative bacterium of the family *Enterobacteriaceae*. Unlike most *Salmonella* species, *S. Typhi* is a human-restricted pathogen, and the clinical syndrome it produces often mimics other acute febrile illnesses, complicating timely diagnosis and management (2). The symptoms of typhoid fever usually include a protracted fever, headache, malaise, diarrhea or constipation, stomach pain, and, occasionally, hepatosplenomegaly. Hematological abnormalities like anemia and thrombocytopenia, as well as biochemical disruptions like changed calcium and potassium levels, can be linked to severe disease and correlate with the severity of the illness and the results of treatment (3,4). Traditional bacterial identification methods, including culture-based, biochemical, and serological assays, remain resource and time-intensive (5), even advanced molecular techniques such as PCR involve laborious experimental procedures, collectively delaying accurate diagnosis and timely treatment of infections (6).

Antimicrobial resistance (AMR), which undermines the effectiveness of antimicrobial treatments and calls for the creation of more effective treatment regimens, is one of the most pressing problems facing worldwide public health (5,7,8). When it comes to patient-level variability and shifting resistance dynamics, clinical decision-making in infectious illnesses usually relies on guidelines generated from surveillance, empirical rule-based techniques like MYCIN, or clinician experience. However, none of these approaches is very flexible (1). Effective clinical management and international control efforts are highly threatened by the advent of multidrug-resistant (MDR) and extensively drug-resistant (XDR) S. Typhi strains, which have drastically reduced therapeutic options for typhoid fever (4,9).

Recent advances in artificial intelligence (AI) and machine learning (ML) have enabled clinical decision support systems (CDSS), which provide real-time personalized antimicrobial recommendations based on patient-specific factors such as laboratory results, comorbidities, and pathogen profiles (6,10). AI is improving epidemiological surveillance, pathogen identification, antimicrobial susceptibility testing, genome sequencing, drug discovery, vaccine development, and phage therapy, in addition to supporting empirical therapy (11). These advancements are changing the overall landscape of managing bacterial infections (11). Better stewardship outcomes, less dependence on broad-spectrum antibiotics, and better alignment with resistance trends are all results of these technologies, especially machine learning and deep learning, which increase the speed, accuracy, and efficiency of clinical decision-making (12,13). ML approaches integrated into CDSS have also shown encouraging results in predicting resistance and optimizing antibiotics as they offer powerful approaches to addressing antimicrobial resistance by integrating diverse datasets, enabling early detection of resistance, streamlining research, and supporting personalized treatment strategies (13).

Counterfactual examination of prescribing decisions is supported by machine learning models that can model treatment outcomes under different drug selections. Applications include using interpretable clustering and fuzzy logic analytics to guide ICU medication and reduce needless antibiotic use in primary care while maintaining efficacy (14,15). The translational potential of probabilistic models in infectious disease management has been highlighted by AI-driven decision support systems like HIV-TRePS, which have shown good predictive accuracy across a variety of patient populations in addition to antibiotic stewardship (16).

Regression of treatment duration and resistance proxy score, while binary classification of suspected typhoid fever and treatment outcomes were the four predictive models for which we applied Extreme Gradient Boosting (XGBoost), an ensemble technique based on gradient-boosted decision trees. It has good performance on diverse clinical datasets, scalability, and computational efficiency (17). In biomedical applications, where missingness, imbalance, and high feature heterogeneity are prevalent, the algorithm optimizes additive tree ensembles using second-order gradient information, incorporates regularization to prevent overfitting, and supports sparse input handling (12). Because of its sophisticated use of parallelization, column subsampling, and shrinking (17), XGBoost exhibits faster training and higher accuracy when compared to conventional gradient boosting frameworks. In precision medicine, where ensemble tree-based learners perform better than linear models in capturing intricate non-linear interactions while maintaining interpretability, these models collectively constitute a multi-task prediction framework that is in line with current trends. Their translational promise in low-resource and high-burden settings is highlighted by recent work that supports their application in clinical outcome predicting, resistance prediction, and infectious disease modeling (18–20).

In this study, we used patient biochemical and demographic data to construct an ML-driven framework for optimizing drug selection in typhoid treatment. Four predictive tasks were specifically addressed by our implementation of Extreme Gradient Boosting (XGBoost): (i) classification of treatment outcomes, (ii) regression of treatment duration, (iii) regression of resistance proxy scores, and (iv) classification of suspected typhoid. In order to compare the actual prescribing decisions with the estimated success probability of antibiotics, we conducted medication simulation trials. This study shows that AI-assisted decision support for typhoid therapy is feasible, and it may find use in precision medicine and antimicrobial stewardship in environments with limited resources.

### 1.1 Clinical and Biological Significance of Dataset Parameters

Understanding the clinical and biological relevance of each parameter in the dataset is crucial for developing an AI model that not only predicts outcomes but also provides interpretable insights aligned with medical knowledge. *Hemoglobin* is a protein in red blood cells responsible for transporting oxygen. Its concentration reflects the blood’s oxygen-carrying capacity. The normal range is 13.5–17.5 g/dL for men; 12.0–15.5 g/dL for women. Hemoglobin levels below these ranges indicate anemia, which can result from various causes, including nutritional deficiencies, chronic diseases, or bone marrow disorders. Anemia in typhoid patients may indicate chronic infection or bone marrow suppression, affecting treatment decisions (21). Blood clotting depends on *platelets*. Bleeding or thrombotic problems may result from abnormal numbers. 150,000–450,000 platelets per microliter of blood (22) is the typical range. Thrombocytopenia, which increases the risk of bleeding, is indicated by counts below 150,000/μL, whereas thrombocytosis, which may indicate inflammation or myeloproliferative diseases, is indicated by levels exceeding 450,000/μL. Typhoid fever frequently causes thrombocytopenia, which can indicate the severity of the illness or its consequences, such as disseminated intravascular coagulation (22). Blood coagulation, muscular contraction, nerve transmission, and bone health all depend on *calcium*. It typically ranges between 8.5 and 10.2 mg/dL. While levels above 10.2 mg/dL indicate hypercalcemia, which may result in kidney stones, bone discomfort, or neurological symptoms (23), levels below 8.5 mg/dL indicate hypocalcemia, which may induce muscle cramps, cardiac arrhythmias, or seizures (24). Due to severe infections or gastrointestinal losses, electrolyte abnormalities, including hypocalcemia, can arise in typhoid patients (25). *Potassium* is essential for preserving cellular activity, especially in muscle and nerve cells. Between 3.5 and 5.0 mmol/L (26) is the typical range. Whereas levels above 5.0 mmol/L indicate hyperkalemia, which can result in potentially fatal cardiac conduction anomalies, levels below 3.5 mmol/L indicate hypokalemia, which causes cardiac arrhythmias, cramping, or muscle weakness. Diarrhea and vomiting in typhoid can lead to hypokalemia, necessitating electrolyte monitoring and replacement (27). The severity of symptoms reflects disease progression and guides treatment intensity. It can be Mild with Low-grade fever, malaise; Moderate with High fever, abdominal pain; and Severe with Delirium, intestinal hemorrhage, or perforation. In typhoid, this correlates with disease stage and complications, influencing hospitalization and treatment strategies (28). Identifying the causative organism confirms the diagnosis and guides antibiotic therapy. *Salmonella enterica serovar Typhi* is the Primary cause of typhoid fever, whereas other Enterobacteriaceae may indicate co-infections or alternative diagnoses. Duration of therapy is determined by disease severity, response to treatment, and presence of complications. Uncomplicated Typhoid may take 7–14 days, while Complicated Cases may require extended therapy. Monitoring treatment duration assesses adherence and effectiveness, influencing outcomes.

## 2. Materials and methods

### 2.1 Code Availability

The source code for all computational work is available through the GitHub repository [https://github.com/SsemuyigaMHC/TyphoidRx] to ensure transparency, reproducibility, and community validation of all procedures.

### 2.2 Libraries and Computational Tools

All analyses, simulations, and predictive modeling procedures were implemented in Python (version 3.12) using a modular set of open-source modules designed for scientific computing and machine learning. In order to effectively organize and modify patient-level clinical and biochemical records, Pandas and NumPy were utilized to handle core data processing and numerical calculations (29,30). Dimensionality reduction, logistic regression, k-fold cross-validation, principal component analysis (PCA), and a range of classification metrics such as accuracy, precision, recall, F1-score, AUC-ROC, and confusion matrix visualizations were carried out using Scikit-learn (31). For ensemble modeling, we employed XGBoost, a highly optimized gradient boosting framework that offers fast and scalable tree-based learning, particularly helpful for tabular and unbalanced clinical data (17). The imbalanced-learn package’s SMOTE (Synthetic Minority Over-sampling Technique) was used to resolve class imbalance in treatment outcome labels. This improved classification robustness by creating synthetic training samples for the minority class (32). Exploratory visualization of high-dimensional patient profiles was done using Advanced data embedding methods, UMAP, and t-SNE (33,34). For model interpretability, model outputs were divided into feature-level contributions based on cooperative game theory utilizing SHAP (Shapley Additive exPlanations), making it possible to thoroughly justify treatment predictions (35). Seaborn, matplotlib-venn, and Matplotlib were used to create the visualizations (36,37). To manage file systems, persist trained models, ensure reproducibility, and track resource utilization during testing, auxiliary Python modules including os, joblib, random, and psutil were employed. This computational framework was executed on a high-performance Linux workstation powered by an Intel Xeon processor and 64 GB RAM, ensuring efficient parallel execution and memory-intensive computation.

### 2.3 Data Acquisition and Pre-Processing

The dataset consisted of 5,760 patient records and 13 columns related to clinical and microbiological data for individuals diagnosed with typhoid fever. The key features included demographic information (*Age* and *Gender*), laboratory parameters (*Hemoglobin*, *Platelet Count*, *Calcium*, and *Potassium*), culture results (*Blood Culture Bacteria* and *Urine Culture Bacteria*), medication administered, treatment duration, and the treatment outcome (Supplementary File 2). The data was subjected to a structured data preprocessing pipeline to clean, explore, enhance, and prepare a clinical dataset for AI-based prediction of typhoid fever treatment outcomes. The dataset was first inspected to verify structure, types, and consistency. Second, missing values were assessed and handled if any, using median imputation for numeric data and ‘Unknown’ for categorical data, reducing bias (38). The outcome class distributions were examined to detect imbalance, a critical insight for classification models. The basic statistical summaries were generated for numeric variables to evaluate central tendency and spread, and distributions were also visualized. The cleaned dataset was ingested and rigorously de-duplicated based on unique patient identifiers. In case of duplicates, only the first record per patient was retained. Records with missing values in the target variables were excluded from further analysis. Microbiological culture results were harmonized by applying label encoding to both *Blood Culture Bacteria* and *Urine Culture Bacteria* fields after replacing nulls with sentinel values.

### 2.4 Feature Engineering

The goal of feature engineering in this analysis was to enhance the predictive value of raw clinical data by encoding, transforming, or combining variables into clinically meaningful features. This process aimed to capture latent patterns associated with typhoid severity, treatment response, and biochemical status, in preparation for further machine learning modeling.

In order to create a severity score for further study, the initial classification of symptoms as Low, Moderate, or High was numerically encoded as 1, 2, and 3, respectively. The agreement between bacterial isolates taken from various biological specimens belonging to the same patient is known as culture concordance. In this study, it specifically denotes whether the same pathogen was isolated from both blood and urine cultures. It often signifies a more severe or invasive disease process, systemic dissemination of the same pathogen, compromised immunity, a higher bacterial load, or delayed clearance (3) and has implications in both diagnostics and treatment selection. Concordance was recorded as 1 if they matched and 0 otherwise. *Salmonella enterica* serovar Typhi (*S. Typhi*) is the primary cause of typhoid fever, a systemic illness transmitted via the fecal-oral route. Diagnosis is ideally confirmed by isolating *S. Typhi* from sterile sites such as blood, bone marrow, or urine, though culture sensitivity can be reduced by prior antibiotics or low bacterial load (3,39). Detection of *S. Typhi* in either blood or urine cultures confirms active infection and informs antimicrobial therapy. The “*Salmonella Presence*” feature flags records where *S. Typhi* appears in either culture field, capturing cases that culture concordance might miss, especially when present in only one sample. *Culture Type* reflects refined culture concordance across systemic compartments. In typhoid fever, *S. Typhi* typically appears in the blood early and may later be detected in urine. Using preprocessed blood and urine culture fields, we engineered a categorical variable as follows: 0 = No growth (both cultures negative/missing), 1 = Positive blood culture only, 2 = Positive urine culture only, 3 = Concordant growth (same species in both), 4 = Discordant growth (different species). This feature captures inter-site infection dynamics without prioritizing *S. Typhi*, enabling broader characterization of culture agreement. *Sex Code* accounts for biological sex, which influences immune function, drug metabolism, and baseline hemoglobin levels. The dataset encoded gender as 0 = Female and 1 = Male; this was directly mapped to the Sex Code variable. Potential therapeutic effects or interactions are captured by medication features. Binary indicators were created for each administered medicine using one-hot encoding of the *Current Medication* column. The *Intensity Score* is a squared severity score that magnifies differences between mild, moderate, and severe symptoms, enabling stronger weighting for highly symptomatic cases. *Normalized Treatment Duration* adjusts raw treatment length by clinical severity to reflect deviations from expected recovery time. It was calculated by dividing treatment duration by the severity score (40). *Resistance Proxy Score* is a composite metric engineered to estimate the likelihood of antimicrobial resistance or treatment failure in suspected typhoid fever cases. It integrates three clinically relevant dimensions: microbiological culture discordance, symptom severity, and treatment duration, all recognized as indicators of poor therapeutic response or resistance (3,4,41,42). Discordant culture results may indicate mixed infections or inadequate empirical antibiotic coverage (4). Prolonged treatment duration often reflects persistent infection or delayed improvement, while higher severity scores capture more complicated disease courses (39,40). Together, these variables reflect known resistance-associated patterns in typhoid. The score was defined as:

Resistance Proxy Score = (1 – Culture Concordance) × Normalized Treatment Duration × Severity Score.

This assigns higher scores to patients with discordant cultures, longer treatments, and more severe symptoms. Concordant culture cases receive a score of 0, reflecting that matched pathogens across sampled sites at baseline indicate a uniform and microbiologically consistent infection pattern, rather than variability arising from mixed or site-specific pathogens. While not a substitute for antimicrobial susceptibility testing, this score serves as a pragmatic surrogate in data-limited settings (3,42). Hemoglobin status was classified as anemic (1), normal (0), or elevated/polycythemia (2) based on sex-specific reference thresholds for biochemical feature engineering; platelet status was coded as thrombocytopenia (1), normal (0), or thrombocytosis (2); calcium status was classified as hypocalcemia (1), normal (0), or hypercalcemia (2); and potassium status was similarly derived as hypokalemia (1), normal (0), or hyperkalemia (2). This allowed raw laboratory values to be converted into clinically interpretable categories for modeling. A derived clinical feature called “suspected typhoid” is used to identify patients who are most likely to have typhoid fever based on a combination of important biochemical indicators, symptom severity, and microbiological evidence. Although definitive diagnosis relies on isolating *Salmonella enterica* serovar Typhi from sterile sites, sensitivity may be reduced due to prior antibiotic use or limited diagnostic capacity in endemic or low-resource settings, resulting in false negatives despite clinical suspicion (39,43). To address this, Suspected Typhoid was engineered to include both confirmed and probable cases. It was computed as a binary variable set to (1) for confirmed *Salmonella* presence in blood or urine cultures, or co-occurrence of moderate to high symptom severity (Severity Score ≥ 2) and biochemical signs of infection specifically, anemia (Hemoglobin Status == 1) or thrombocytopenia (Platelet Status == 1), and 0 otherwise. These indicators have been previously associated with typhoid pathophysiology and reflect the systemic impact of the disease (44). By integrating clinical and laboratory signals, this feature enhances diagnostic sensitivity for identifying probable typhoid cases, especially where culture confirmation is unavailable. The summary is shown in Table S1.

### 2.5 Metabolomic Simulation

Metabolomic simulation, as used in this work, is the computational synthesis of metabolite-related features utilizing clinical and biochemical data to mimic possible metabolic changes in typhoid fever patients. This method supports personalized medicine by collecting host-pathogen metabolic interactions that may impact treatment results or resistance patterns, improves predictive power, and makes multi-omics integration easier by connecting biochemical markers with metabolomics-derived phenotypes. We used a hybrid approach that blends rule-based simulation with machine learning-driven latent profiling to mimic metabolomic layers without the need for direct metabolomic data. Without depending on pre-established pathway linkages or external databases like KEGG, which might not be unique to typhoid fever, our approach provides biological interpretability and catches intricate data patterns. It also lessens the possibility of overfitting that comes with a generative AI model.

#### 2.5.1 Rule-Based Simulation

By mapping variations in biochemical markers to possible metabolic disturbances, the rule-based simulation used regular clinical biochemistry data to infer metabolic disruptions. Six features at the binary metabolite level were simulated: *SimMet_EnergyDisruption*: Marked as 1 if hemoglobin levels were less than 12 g/dL for females or less than 13 g/dL for males, indicating that ATP synthesis was impacted by poor oxygen delivery. If the platelet count was greater than 450,000/μL, *SimMet_InflammatoryStress* was flagged as 1, indicating that systemic inflammation changes metabolic fluxes and cytokine signaling. In *SimMet_MembraneInstability*, calcium values <8.5 or >10.5 mg/dL were flagged as 1 (45). If potassium levels were less than 3.5 or greater than 5.0 mmol/L, *SimMet_ElectrolyteDisruption* was flagged as 1, indicating disruptions in cellular metabolism and acid-base balance. If the severity score was ≥2, *SimMet_CatabolicBurst* was flagged as 1, indicating elevated protein and energy catabolism linked to systemic inflammation (46). *SimMet_HypoxiaStress:* Marked as 1 in cases of low hemoglobin levels, this indicates a decreased ability to carry oxygen, which leads to compensatory metabolic adjustments like anaerobic glycolysis (47) and 0 the other way round for all the features. When direct metabolomics profiling is not practical, these features help generate hypotheses, stratify risks, and interpret disease-related states functionally by approximating actual metabolic phenotypes.

#### 2.5.2 Unsupervised Latent Metabolite Feature Extraction

To extract latent metabolomic features, we applied PCA to a specific set of metabolic data, including clinical biomarkers and indicators of simulated metabolic stress. Before using PCA, all features were standardized using z-score normalization to ensure equal weighting across variables. Two components (n_components=2) were used in PCA to capture the main axes of variation in the combined clinical and simulated metabolomics profile. The generated principal components (PC1 and PC2) were used as input features in prediction models and functioned as compressed, latent metabolic fingerprints. This strategy is in line with well-established metabolomics techniques, where dimensionality reduction improves prediction accuracy and model generalizability by distilling physiologically significant variance into interpretable components (48,49)(50).

### 2.6 Model Training and Optimization

To evaluate predictive performance across clinically relevant outcomes, we developed and optimized four supervised machine learning models using the whole dataset: (i) a binary classifier for treatment outcome, (ii) a regression model for treatment duration, (iii) a regression model for resistance proxy score, and (iv) a classifier for suspected typhoid diagnosis. All models were implemented using the Extreme Gradient Boosting (XGBoost) algorithm, known for its robustness in structured healthcare data and superior performance in tabular domains (17). For all models, before training, Features with possible leakage and high multicollinearity (based on Pearson correlation analysis) were removed. All Boolean variables were encoded. Hyperparameter tuning was performed using RandomizedSearchCV across 30 sampled configurations, evaluating combinations of learning rate, tree depth, regularization parameters, and sampling ratios, improving model performance and avoiding overfitting (51). The search space included: n_estimators ∈ {100–500}, max_depth ∈ {3–10}, learning_rate ∈ [0.01–0.3], subsample ∈ [0.5–1.0], colsample_bytree ∈ [0.5–1.0], and reg_lambda ∈ [0–10]. All models were trained using five-fold cross-validation with random shuffling and stratification where applicable. Hyperparameter tuning was executed within the same cross-validation folds used for model evaluation, ensuring that performance metrics reflected only the optimized configuration without introducing external information. The best-scoring configuration from this procedure was refit on the full training data of each seed. For classification tasks, SMOTE was applied to address class imbalance (32) and cross-validation was done using ROC_AUC. SMOTE was applied only to the training partition within each cross-validation split to correct class imbalance in the treatment-outcome and infection-classification tasks. It was implemented using the default k-nearest-neighbor setting (*k* = 5) and the standard oversampling strategy that increases the minority class to match the majority class within each fold. Importantly, SMOTE was *not* applied to the held-out validation splits, thereby preventing information leakage and ensuring unbiased performance estimation. For regression tasks, the average cross-validated R^2^ was used to choose the final model configuration.

### 2.7 Model Evaluation

For each model, unbiased performance estimates were obtained using a repeated nested cross-validation (nested-CV) framework ma(52). To completely separate hyperparameter tuning from performance assessment and prevent optimistic bias during model selection, Nested-CV was selected. 10 separate random seeds (100–109) were used to repeat the whole nested-CV pipeline in order to account for stochastic variation in both data splitting and model initialization. A brand-new nested-CV procedure, replete with new random splits for the inner and outer cross-validation loops, was run for every seed. Each seed functioned as a separate replication of the whole tuning-and-evaluation process; no models, results, or hyperparameters were exchanged among seeds.

A 5-fold outer cross-validation cycle produced objective evaluations of predicted performance for every seed. A 5-fold inner cross-validation loop was utilized for RandomizedSearchCV hyperparameter adjustment within each outer-fold training split (53). The hyperparameter search space, which varied in tree depth, learning rate, subsampling ratios, column sampling ratios, number of estimators, gamma regularization, and L2-regularization (λ), was the same for all models. For each inner loop, thirty different random hyperparameter combinations were assessed. The outer loop was strictly reserved for final performance grading, while model selection took place only within the inner loop.

The main optimization measures for XGBoost regressors were mean absolute error (MAE) and coefficient of determination (R²) (54). The area under the receiver operating characteristic curve (ROC AUC) was used to assess model discrimination for XGBoost classifiers. The unbiased assessment of model performance for each seed-level nested-CV run was represented by the mean of the outer-fold scores. Random initialization, random CV partitioning, and stochasticity in the randomized hyperparameter search all contribute to the variability in the final reported performance for each prediction task, which is expressed as mean ± standard deviation among the ten seeds. Repetitive seeds were only used for variance estimates and robustness evaluation; neither ensemble averaging nor model aggregation were carried out (55,56).

### 2.8 Drug Recommendation Simulation

To evaluate the potential for improved treatment outcomes via drug switching, we simulated counterfactual drug assignments for each patient in the test set using the trained treatment outcome model. The simulation aimed to estimate which of three candidate antibiotics, amoxicillin, azithromycin, or ceftriaxone, would maximize the predicted probability of treatment success for each individual, assuming all other clinical variables remained constant. Each patient’s engineered feature vector was replicated three times and manipulated to simulate the administration of each possible antibiotic. Medication features were encoded in a mutually exclusive fashion to reflect realistic prescribing practice. All features were passed through the outcome classifier, which returned predicted probabilities of treatment success. The top two ranked antibiotics (by predicted success probability) were retained for analysis. The resultant enriched dataset allowed for retrospective comparison between real-world prescribing and model-recommended alternatives. The resultant predictions were subjected to further analysis.

## 3.0 Results and discussion

### 3.1 Dataset Overview and Loading

The class distribution is shown in Supplementary Figure S1. The raw dataset (N = 5,760 records, 13 variables) contained missing values in several fields: *Symptoms Severity* (n = 629), *Blood Culture Bacteria* (n = 832), *Treatment Duration* (n = 731), and *Treatment Outcome* (n = 576) (Supplementary File 2, Sheet: Missing Report). After listwise deletion of records with unresolved outcome variables, the final analytical cohort included 5,184 complete records. The *Treatment Outcome* variable exhibited moderate imbalance, with 2,810 cases (54.2%) classified as *Successful* and 2,374 (45.8%) as *Unsuccessful*. This ∼1.18:1 ratio, although not severe, may bias classifiers toward the majority class. To mitigate this, SMOTE was scheduled for use during model training stage (32).

The demographic characteristics of the study cohort indicated a nearly balanced sex distribution, with males comprising 51.9% of participants. The mean age was 43.1 years (SD = 14.94; range: 18–70 years). Laboratory and clinical biomarker assessments showed a mean hemoglobin concentration of 13.0 g/dL (SD = 2.65), with values in the lower quartile falling below standard reference thresholds, suggesting that a subset of participants may have exhibited subclinical anemia. The mean platelet count was 209,115/µL (SD ≈ 50,000), which remained within physiological reference ranges. Serum electrolyte analysis revealed mean calcium and potassium concentrations of 9.06 mg/dL and 4.56 mmol/L, respectively. The average duration of treatment was consistent with standard therapeutic regimens, with a mean of 9.98 days (SD = 2.90) (Supplementary File2, Sheet: Statistics).

The categorical variables followed distributions that made clinical sense in the context of typhoid fever. Symptom severity was evenly split across the three categories: Low (33.3%), Moderate (33.3%), and High (33.3%), capturing the full spectrum of disease presentation observed in practice. Blood culture results showed three dominant organisms: *Staphylococcus* spp. (29.7%), *Salmonella typhi* (≈29.7%) and *Escherichia coli* (≈22.5%). The presence of *S. typhi* reflects its role as the primary cause of enteric fever, while the frequent isolation of *Staphylococcus* and *E. coli* may point to co-infections or diagnostic overlap with other febrile illnesses, a known challenge in endemic settings where clinical symptoms are often nonspecific (3,4). Urine culture results were dominated by *E. coli* (35.9%), which aligns with its established role as the most common urinary tract pathogen. Within a typhoid dataset, this finding also highlights the possibility of concurrent infections that can complicate treatment response and prolong recovery. Patterns of prescribed medication reflected current treatment realities. Amoxicillin (35.8%) was the most frequently recorded drug, followed by Ceftriaxone (30.3%) and Azithromycin (29.9%). This mirrors global and regional practice, where amoxicillin continues to be used despite declining efficacy, ceftriaxone is often reserved for hospitalized or severe cases, and azithromycin has gained ground as a convenient oral option with reliable activity against multidrug-resistant *S. typhi* (41). The predominance of these three antibiotics in the dataset underscores their frontline status in typhoid therapy, but also reflects the growing tension between clinical necessity and rising antimicrobial resistance (Figure S1).

To evaluate inter-feature relationships and assess the risk of multicollinearity in downstream modeling, a Pearson correlation heatmap was generated for all numeric variables (Figure S2). As expected, diagonal entries showed perfect self-correlation (r = 1.0). Pairwise correlations between distinct features were uniformly weak, with no coefficient exceeding |r| = 0.06. The strongest positive correlation, albeit negligible, was observed between Treatment Outcome and Platelet Count (r = 0.036), followed by Treatment Duration and Treatment Outcome (r = 0.020). Age displayed a minor inverse association with Treatment Duration (r = –0.059), suggesting that older patients may have slightly shorter treatment courses. Interestingly, electrolyte values such as calcium and potassium showed an almost zero correlation (r = 0.0016), which diverges from expected physiological co-regulation and may reflect either biological heterogeneity or measurement noise within the dataset. Taken together, the correlation matrix confirmed the absence of multicollinearity, with all inter-variable relationships close to zero. This finding supports the application of non-linear machine learning algorithms such as XGBoost or Random Forest, which are robust to independent feature contributions. Moreover, the lack of strong correlations indicates minimal redundancy among predictors, thereby preserving their individual discriminative value for multivariate modeling.

### 3.2 Feature engineering

Robust modeling of typhoid-related clinical variability was made possible by the conversion of unprocessed clinical, biochemical, and microbiological information into analytically tractable and physiologically significant artificial characteristics. These manufactured features consistently displayed patterns that matched their intended semantic definitions, as seen in Figure S3. There was no discernible imbalance or propagation of missing data, and key category features maintained logical distributions (43). Clinical variety among the patient population was shown by the multimodal or right-skewed distributions of continuous designed variables (39,57,58).

#### 3.2.1 Simulated Metabolomic Profile Distributions

The six features that were produced showed different distribution patterns, as seen in Figure S3. SimMet_EnergyDisruption and SimMet_HypoxiaStress were common, indicating anemia that is frequently seen in systemic infections, such as typhoid (59). In line with cytokine-driven platelet increases documented in typhoid fever (60), SimMet_InflammatoryStress identified a subpopulation with increased inflammatory activity. SimMet_ElectrolyteDisruption and SimMet_MembraneInstability were less common (4). In the absence of direct metabolomics testing, these characteristics provided significant approximations of underlying metabolic abnormalities and were both clinically interpretable and in line with recognized biochemical thresholds.

#### 3.2.2 Dimensionality Reduction and Latent Biochemical Signatures

Using PCA on both raw biochemical markers and simulated metabolomics features, two notable latent components (PC1 and PC2) were found (Figure 2). A systemic illness load was indicated by PC1’s weighting primarily on anemia and inflammatory signs, but PC2’s recording of fluctuations in membrane and electrolyte stability suggested renal or mitochondrial involvement. Orthogonal tendencies were noted by the constituent parts. The dimensionality reduction preserved significant biological structure, as seen by the grouping of suspected or confirmed typhoid patients along particular PC1-PC2 space regions. This approach mirrors conventional metabolomics workflows where PCA is used to compress high-dimensional data while maintaining key biological variance (48,61). More precise modeling was made possible by this extensive feature set.

**Figure 1:**
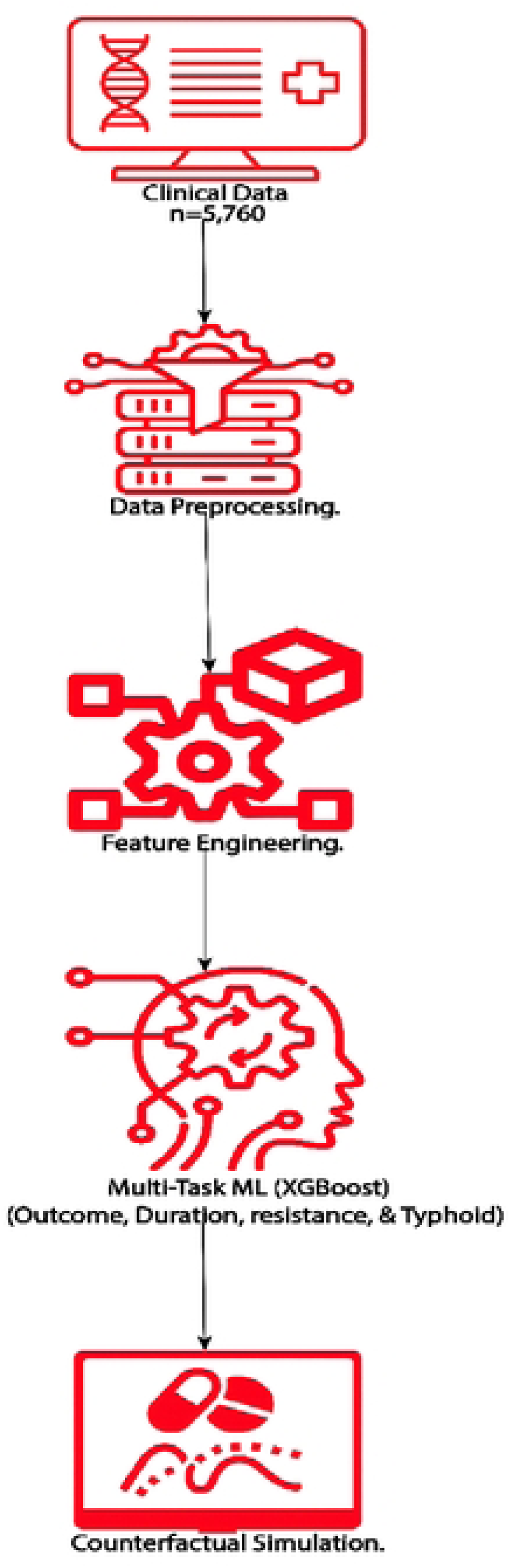
End-to-end study workflow. Clinical and laboratory data were pre-processed and transformed into engineered features, used to train multi-task XGBoost models, and a counterfactual drug-Simulation module estimated patient-specific alternative antibiotic options.

**Figure 2:**
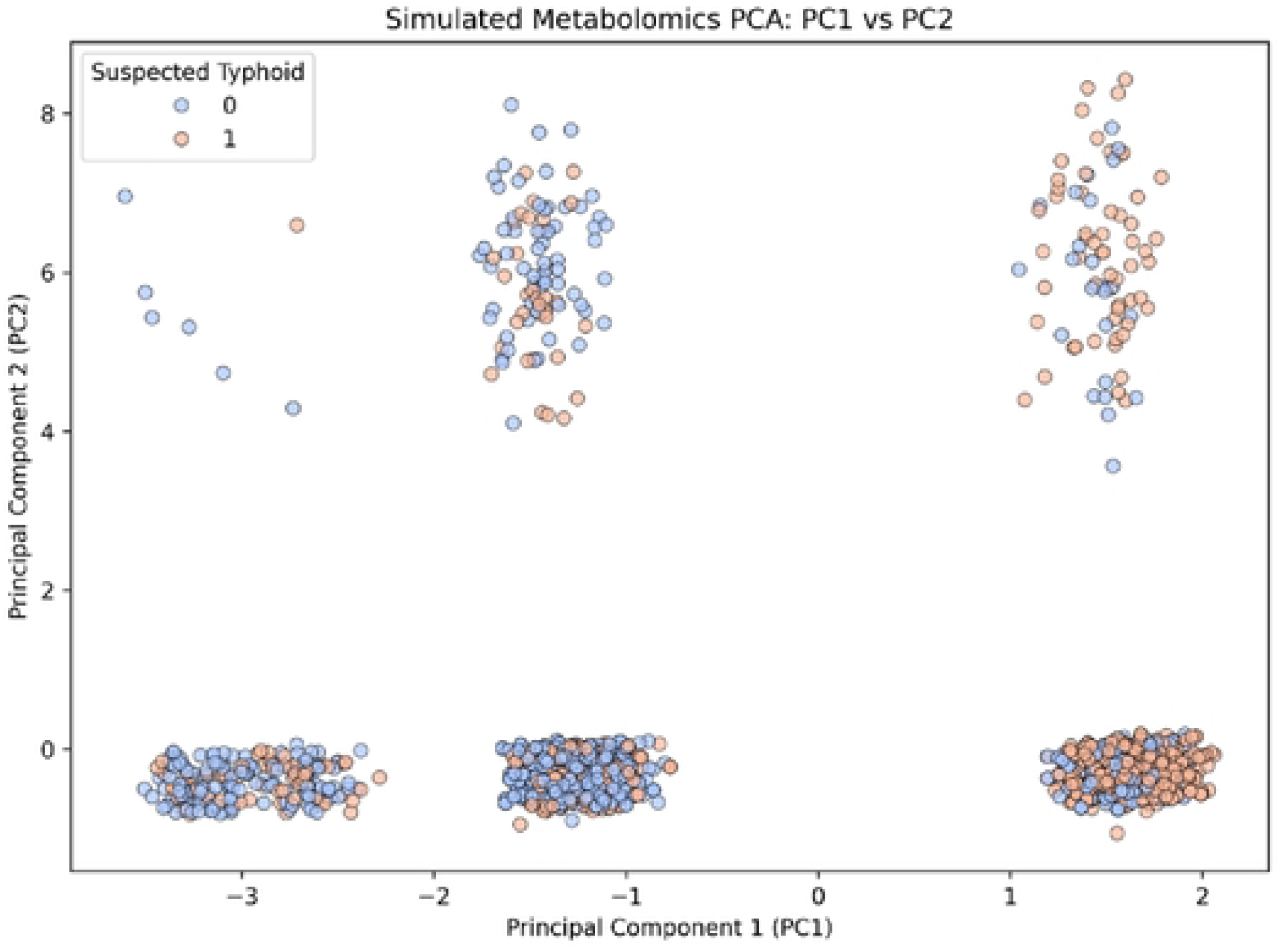
PCA projection of metabolomic and biochemical feature space.

### 3.3 Model Overview and Training Performance

The final set of prediction models was developed using a 75:25 training set to test set ratio using clinical datasets that had been carefully engineered, deduplicated, and cleaned. A customized feature set for each model was used as input features. The machine learning framework used stratified 5-fold cross-validation over 30 hyperparameter configurations per model and grid search to achieve 150 fits per task. In clinical prediction tasks, XGBoost was used as the basis estimator because of its effectiveness and scalability (17).

#### 3.3.1 Treatment Outcome Model

The model was trained on data from 3,480 patients to predict binary treatment outcomes after 408 rows with missing severity values were removed. Up sampling the original dataset indicated a considerable class imbalance (success: failure ∼ 1.2:1), and created a balanced training set (n = 1,895 per class). Laboratory results and metabolomic simulations were among the 21 features that were used in the final model. With a cross-validated ROC AUC of 0.9993, the model demonstrated near-perfect discrimination. The best configuration used n_estimators=300, max_depth=6, learning_rate=0.05, subsample=0.8, reg_lambda=0.1, and colsample_bytree=1.0 (62).

#### 3.3.2 Treatment Duration Model

The regression model for predicting treatment duration was trained using 22 selected features. Variance Inflation Factor (VIF) analysis was used to eliminate multicollinearity, and known leakage variables were excluded. After cross-validation, the final model achieved an R² of 0.7985, indicating strong predictive performance. The optimal model used n_estimators=300, max_depth=8, learning_rate=0.1, subsample=0.9, colsample_bytree=0.7, and reg_lambda=10 (3).

#### 3.3.3 Resistance Proxy Model

The final model was trained on 3,480 clean observations after excluding 408 rows with missing score components. The feature set (n = 20) included antimicrobial exposure, lab metrics, and simulated metabolomic features. The model achieved a cross-validated R² of 0.9020, confirming a strong approximation of resistance likelihood using engineered proxies. The best parameters were n_estimators=300, max_depth=8, learning_rate=0.1, subsample=0.9, colsample_bytree=0.7, and reg_lambda=10 (39,57).

#### 3.3.4 Suspected Typhoid Model

To identify instances of clinically suspected typhoid, an approximately balanced dataset (n = 3,888; 1971 positive, 1917 negative) was used with 20 features spanning clinical, biochemical, and simulated metabolomic attributes. The model showed strong discrimination with a cross-validated ROC AUC of 0.9853. The final hyperparameters were n_estimators=300, max_depth=10, learning_rate=0.1, subsample=0.9, colsample_bytree=0.7, and reg_lambda=10 (59).

### 3.4 Test Set Evaluation and Generalization

The performance of all models was thoroughly assessed across ten separate runs using a bootstrap-based resampling technique on a held-out test set of 1296. This evaluation strategy enhances robustness and quantifies generalization error by incorporating variability introduced by random sampling (63).

#### 3.4.1 Treatment Outcome Classification

With an average ROC AUC of 0.9998 ± 0.0001, the outcome classifier demonstrated remarkable generalization (Table 1 and Figure 3A). On the test set (n=1,296), the total classification accuracy was 99%, and both positive and negative classifications performed equally (precision = 0.99; recall = 0.99–1.00). Strong stability is indicated by the macro-averaged F1-score of 0.99 and the low variance across bootstrap folds (Table 2). These results bolster the clinical outcome prediction’s external validity and robustness. Reliable class identification and a clear predictive structure in the feature space are suggested by high ROC AUC values (Figure 3A). The confusion matrix (Figure S4A) demonstrates near-perfect concordance between observed and predicted labels, with true negative (n = 589) and true positive (n = 699) predictions much outnumbering incorrect predictions (false negatives = 3, false positives = 5).

**Figure 3:**
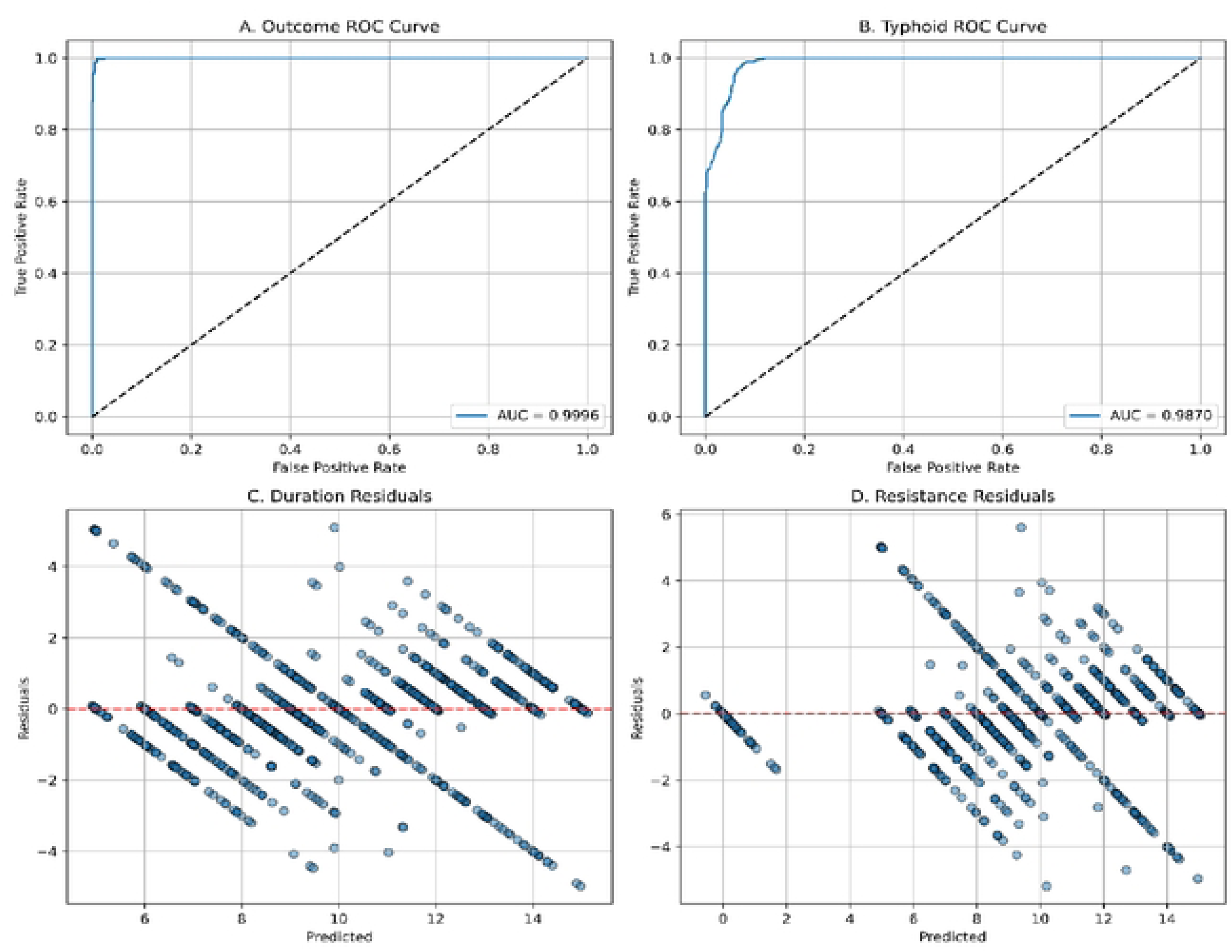
ROC and Residual Plots for Classification and Regression Models. (A) and (B) show ROC curves for the outcome and suspected typhoid classifiers, respectively, indicating strong discriminative ability. (C) and (D) display residual plots for treatment duration and resistance proxy regression models, both showing tight error distributions without systematic deviation.

**Table 1:** Evaluation metrics for all four predictive models on the held-out test set using 10-fold bootstrap resampling. Regression models report R², MAE, MSE, and RMSE; classification models report ROC AUC scores. All models show excellent generalization performance

| Task | Metric | Mean ± SD |
| --- | --- | --- |
| **Treatment Duration (regression)** | $R^2$ | $0.8027 \pm 0.0054$ |
| | MAE | $0.7841 \pm 0.0324$ |
|  | MSE | 1.6423 |
|  | RMSE | 1.2815 |
| **Resistance Proxy (regression)** | $R^2$ | $0.9014 \pm 0.0036$ |
| | MAE | $0.7596 \pm 0.00334$ |
|  | MSE | 1.5922 |
|  | RMSE | 1.2618 |
| **Treatment Outcome (classification)** | ROC AUC | $0.9996 \pm 0.0000$ |
| **Suspected Typhoid (classification)** | ROC AUC | $0.9870 \pm 0.0000$ |

**Table 2:**
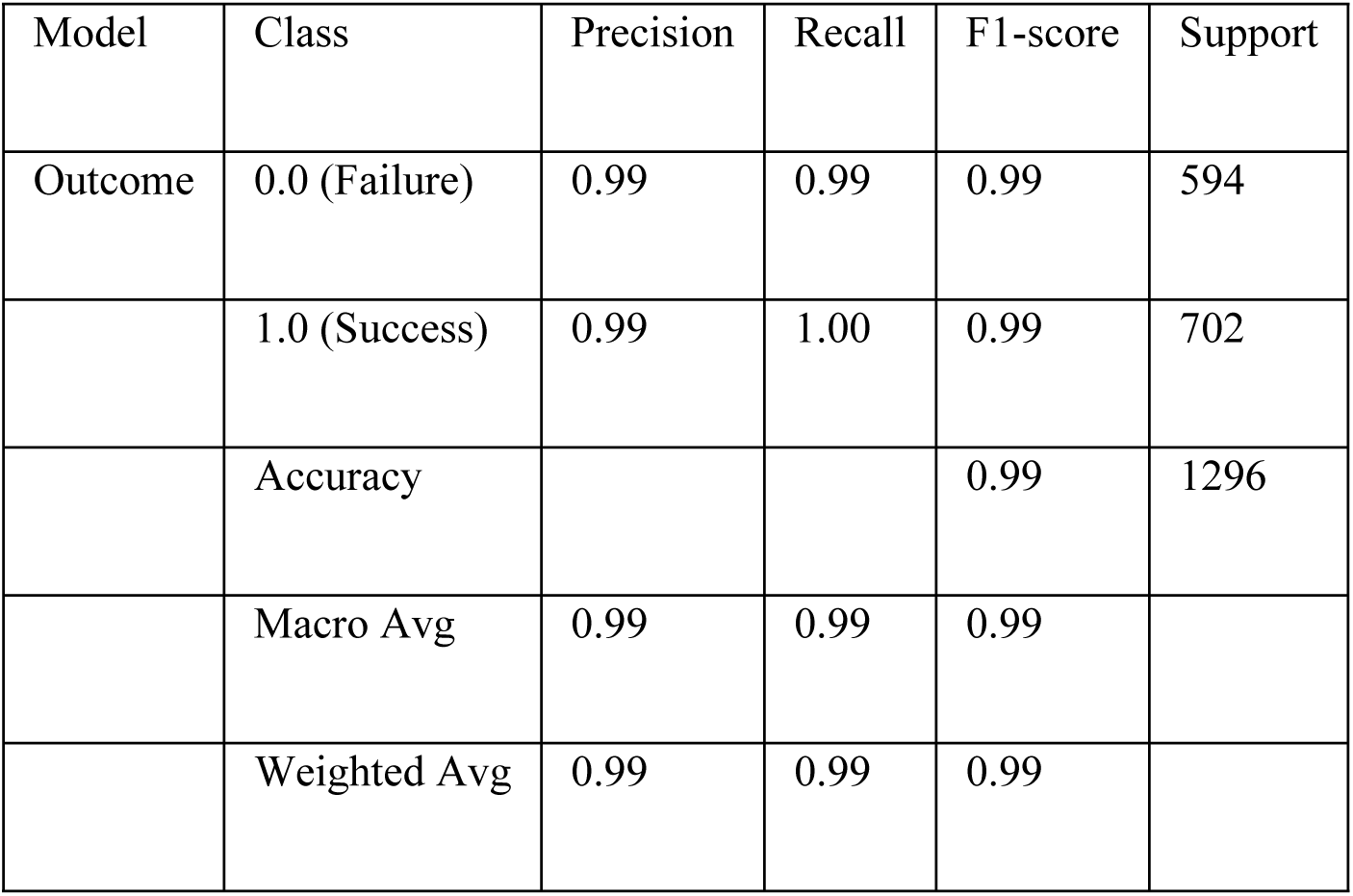
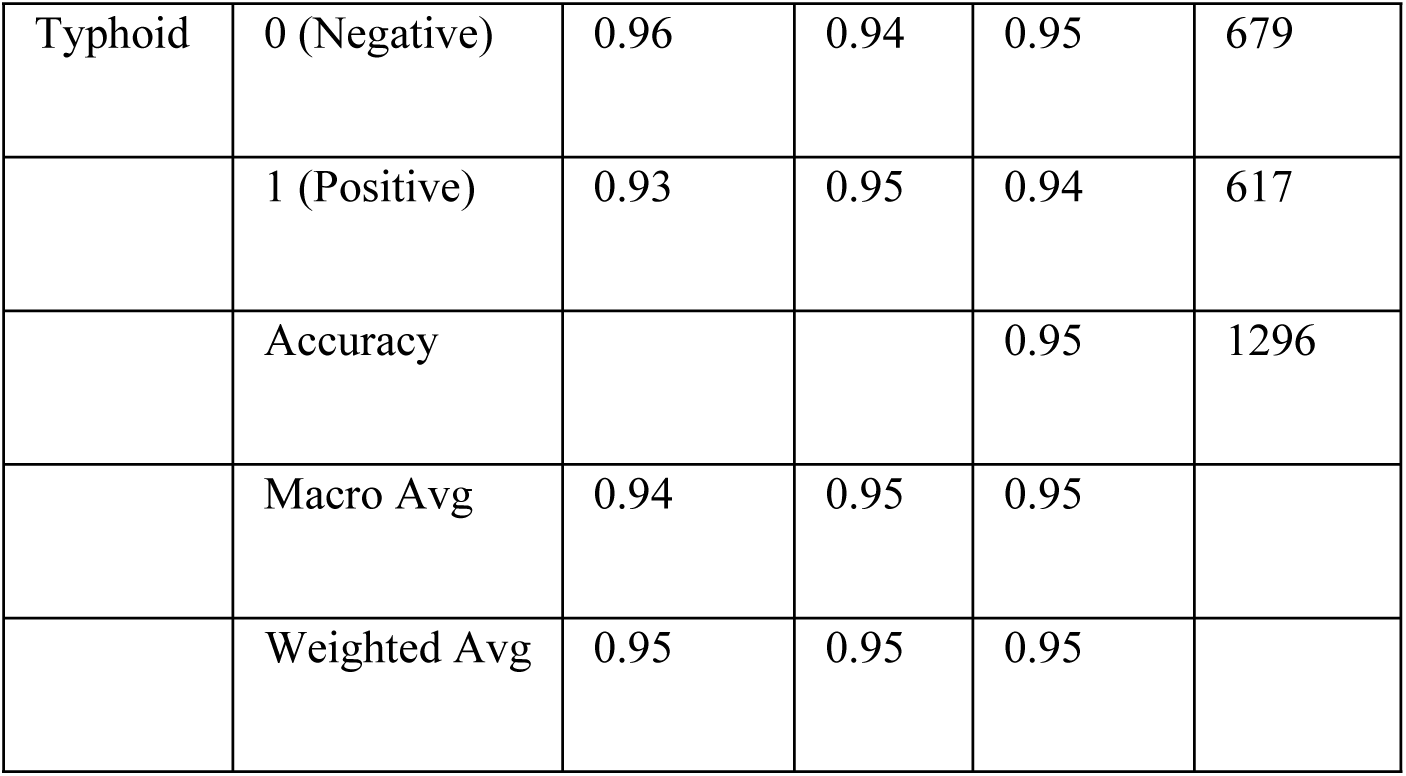
Precision, recall, F1-score, and accuracy for binary classification tasks on the test set. The outcome model performs slightly better overall, while the typhoid model maintains balanced class-wise metrics.

#### 3.4.2 Treatment Duration Regression

The treatment duration model achieved perfect generalization on the test set, with an average R² of 0.8027 ± 0.0054 and a mean absolute error (MAE) of 0.7841 ± 0.0324 days (MSE: 1.6423, RMSE: 1.2815) (Table 1). Model reproducibility across bootstrap iterations and consistent results across all diagnostic plots validate its robustness. This performance likely stems from the high fidelity of numerically cleaned duration labels and biologically meaningful predictors (64). Duration residuals (Figure 3C) were narrowly distributed around zero with no apparent systematic bias, consistent with well-calibrated continuous regression. The evenly scattered points across predicted values demonstrate that error variance was largely stable.

#### 3.4.3 Resistance Proxy Regression

The resistance proxy model also demonstrated outstanding generalization performance, with an average R² of 0.9014 ± 0.0036 and MAE of 0.7596 ± 0.0334. Despite the engineered nature of the target, evaluation on unseen data yielded low error, as confirmed by regression diagnostics (MSE: 1.5922, RMSE: 1.2618). Although proxy variables are not substitutes for lab-confirmed antimicrobial resistance, their strong predictive performance reinforces their value in low-resource simulation, risk stratification, and early warning systems (65). Resistance residuals (Figure 3D) displayed wider dispersion and clustered deviations, particularly at higher predicted values. Such patterns suggest that while the resistance model captures broad outcome tendencies, additional explanatory variables such as pathogen-specific genomic features or prior antimicrobial exposure history may be required to reduce unexplained variance.

#### 3.4.4 Suspected Typhoid Classification

The overall accuracy of the suspected typhoid classifier was 95%, with a robust average ROC AUC of 0.9834 ± 0.0007 (Table 1 and Figure 3B). Positive and negative class-specific F1-scores were 0.94 and 0.95, respectively, and macro-averaged precision and recall were continuously higher than 0.93 (Table 2). Such model can offer critical utility in endemic areas (4,57). The legitimacy of the constructed pipeline and the viability of proxy-enhanced modeling in infectious disease contexts were supported by the remarkable stability and generalizability of all models throughout numerous bootstrap runs. The confusion matrix (Figure S4B) shows that the typhoid classification model achieved high accuracy, correctly identifying 638 negative cases and 587 positives, with only a limited number of false positives (n = 41) and false negatives (n = 30). The low error rates across both classification models underscore their reliability for binary clinical classification tasks (8). The evaluation indicates that both outcome and typhoid classifiers achieve robust categorical performance with negligible misclassification.

It is important to note that the dataset originally comprised only patients who were clinically managed as typhoid-positive cases. Consequently, the negative class used in model training and evaluation was derived through feature engineering rather than direct clinical annotation. Specifically, patients who did not satisfy the composite “suspected typhoid” criteria were designated as negatives, despite being considered positive by clinicians in routine practice. This distinction underscores that the classifier’s performance reflects separation between “probable typhoid” and “less consistent typhoid presentations,” rather than true positives versus true negatives in the strict diagnostic sense. While this limits direct comparability with gold-standard microbiological definitions, it also highlights an important contribution: the model identifies internal structure within a uniformly positive dataset, effectively distinguishing subsets of patients who may be over-classified as typhoid in frontline practice. Moreover, by producing probabilities rather than absolute labels, the classifier provides a spectrum of likelihood, offering a more nuanced view of case heterogeneity than the binary clinical approach. In this way, the approach demonstrates how computational methods can reveal hidden diagnostic heterogeneity and potentially refine clinical case definitions in endemic settings.

### 3.5 Model Calibration and Probability Reliability Assessment

When making decisions at the individual or community level, calibration establishes how reliable the anticipated probability is (66). In order to evaluate the dependability of probabilistic results from predictive models, we used scikit-learn’s calibration curve analysis (67) to create a post hoc calibration analysis. For each model, predicted class probabilities were generated, and calibration curves were constructed using 10 equal-width bins. These curves plot the mean predicted probability against the observed fraction of positive outcomes. A reference line (y = x) was included to indicate perfect calibration, where predicted probabilities align exactly with empirical frequencies. Deviations from this diagonal reflect over– or under-confidence in model outputs. In addition, the Brier score, a proper scoring rule for probabilistic predictions, was computed to quantify overall calibration quality (68).

The Outcome Model’s exceptionally low Brier score (0.0110) indicates that there is little difference between expected and actual results, which is in line with clinical model best-practice guidelines (69). This calibration level shows proper uncertainty quantification in addition to strong discrimination (Figure 4A). With a Brier score of 0.0432, despite being less calibrated than the outcome model, the Typhoid Model (Figure 4B) nonetheless performs well enough for many public health applications and stays under acceptable calibration error limits (67).

**Figure 4:**
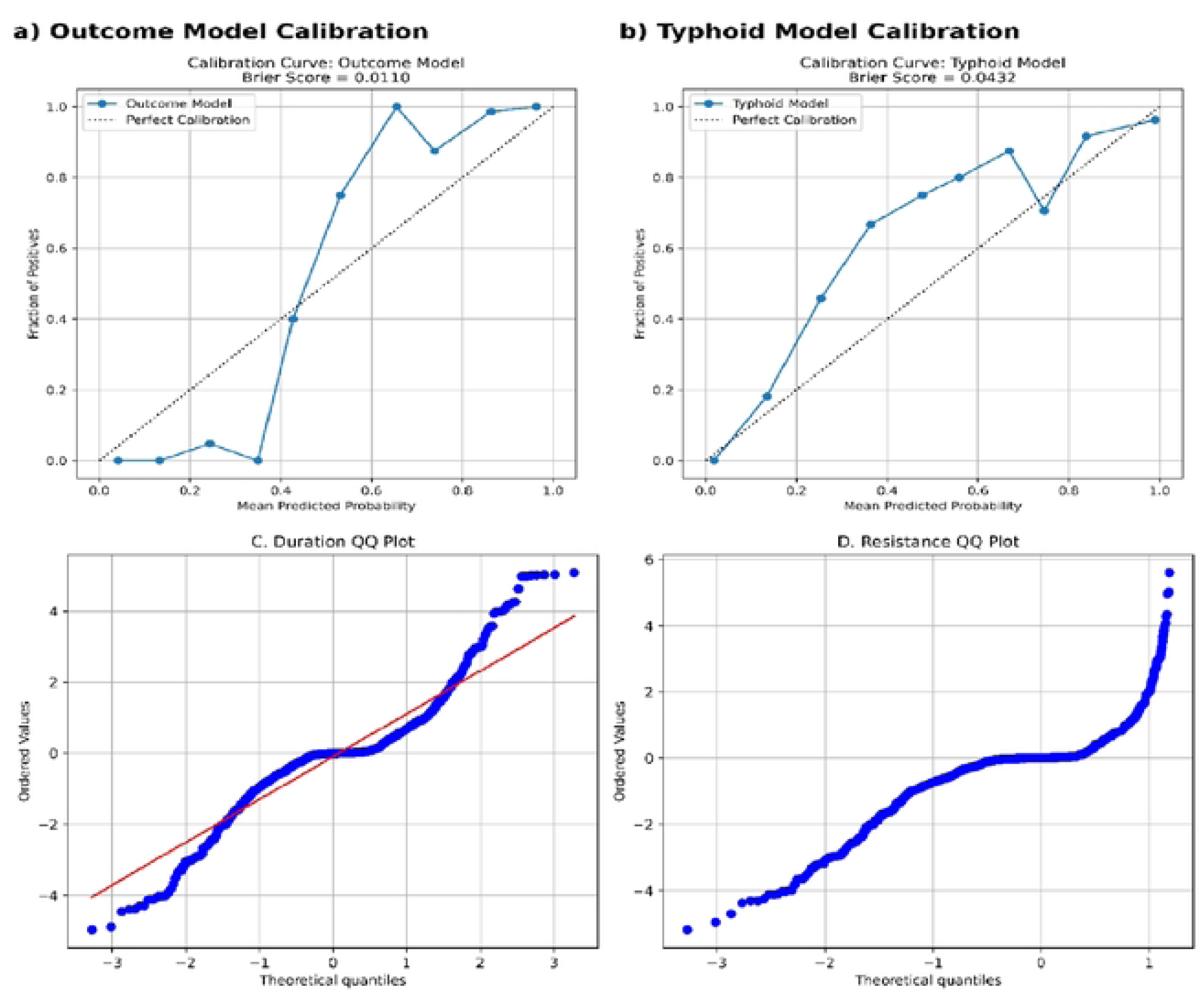
Calibration and diagnostic plots of predictive models. The Treatment Outcome model (a) demonstrated near-perfect calibration, while the Suspected Typhoid model (b) showed good overall calibration with slight overestimation at higher probabilities. The dashed diagonal in both panels represents perfect calibration. Quantile-Quantile plots for treatment duration (c) indicated approximate normality with minor tail deviations, whereas resistance predictions (d) displayed marked departures from normality with heavier tails, reflecting unmodeled heterogeneity in resistance outcomes.

Model calibration for continuous predictions was assessed via Q–Q plots (Figure 4C–D). For treatment duration, observed values aligned closely with theoretical quantiles along the reference line, indicating approximate normality in error distribution. Slight deviations at the tails suggest minor heteroskedasticity, but overall, the pattern supports the model’s adequacy in capturing variance structure. In contrast, resistance predictions exhibited stronger departures from linearity, particularly in the upper quantiles. This curvature points to systematic deviation from normal residual assumptions, potentially reflecting unmodeled heterogeneity in resistance determinants. These findings highlight both the strengths and areas for refinement, particularly the need to integrate richer biological or clinical covariates when modeling resistance outcomes.

### 3.6 Model Explainability and SHAP-Based Patient Stratification

To enhance the interpretability of the trained machine learning models, SHAP analysis was performed (70). For each task, the corresponding XGBoost model (17) and its specific test feature set were loaded.

Features were aligned and padded where necessary to ensure compatibility with the trained model. SHAP values were computed using TreeExplainer and optimized for gradient-boosted tree ensembles. The top 20 features were extracted based on mean absolute SHAP values and visualized using beeswarm plots to summarize both global and individual feature contributions (70). To examine feature consistency across predictive tasks, overlaps in top features were quantified and visualized using a Venn diagram and pairwise heatmap (71). To assess patient-level similarity in model explanations, SHAP value matrices across the four tasks were concatenated per patient. The combined matrix was standardized using z-score normalization and clustered using KMeans (k=5). Dimensionality reduction was performed via Uniform Manifold Approximation and Projection (UMAP) (33) and t-distributed Stochastic Neighbor Embedding (t-SNE) to visualize latent patient subgroups based on SHAP attribution signatures.

#### 3.6.1 Global Feature Importance Across Models

SHAP analysis shows that each of the four prediction models has model-specific yet biologically important feature importance patterns (Figure 5, Table S2). The treatment outcome model’s primary predictors were platelet count, age, hemoglobin (g/dL), potassium (mmol/L), and calcium (mg/dL), highlighting the significance of hematologic and biochemical health in assessing treatment efficacy. Notably, Severity_Score and Culture_Type also contributed significantly, supporting the idea that clinical severity and pathogen profile affect therapy response (4,72). The top predictors for the treatment duration regression model were still age, potassium, platelet count, and hemoglobin (g/dL). Treatment outcome, severity score, and culture type also had an impact, suggesting that both clinical and physiological characteristics are linked to longer recovery times (41).

**Figure 5:**
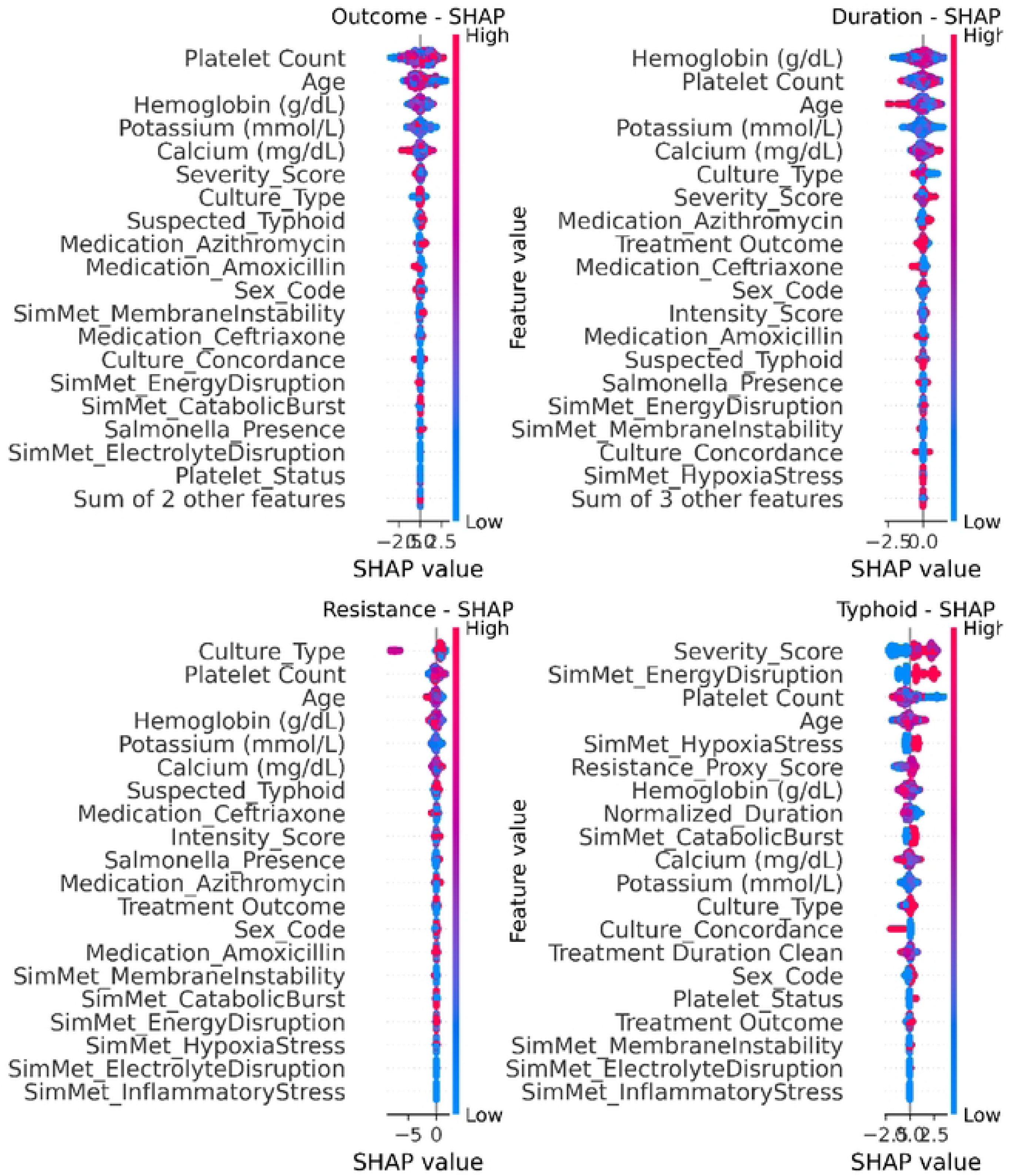
SHAP beeswarm plots for the four predictive models. Each plot displays the top 20 most influential features based on their SHAP values, which reflect their impact on the model’s predictions. Color gradients represent the original feature values.

Platelet count and culture_type were the main drivers of the resistance score regression model, followed by age, hemoglobin, and potassium. The mechanistic significance of microbial physiology in resistance patterns is shown by the appearance of several simulated metabolic stress traits, including SimMet_EnergyDisruption and SimMet_MembraneInstability. These findings support established relationships between antibiotic resistance and culture discordance. Lastly, Severity_Score was the most powerful driver in the suspected typhoid classification model, followed by SimMet_EnergyDisruption, Age, and Platelet Count. In line with research on typhoid-related cytopenias, the existence of Resistance_Proxy_Score and hemoglobin further suggests that systemic stress, inflammation, and hematologic disruption are important diagnostic markers in enteric fever.

#### 3.6.2 Cross-Model Feature Overlap and Modularity

The Venn diagram demonstrates substantial overlap in the top-ranked predictors among the Outcome, Duration, and Resistance models, with 16 features shared across all three. Notably, Typhoid showed relatively less intersection, sharing only 14–15 features with the others, consistent with its task-specific target. Despite this convergence, each model also retained unique predictive signals. SimMet_EnergyDisruption and SimMet_CatabolicBurst showed pronounced importance in Typhoid prediction, reflecting more specialized metabolic stress markers likely associated with infection severity. The pairwise overlap heatmap further quantifies this pattern, and this reflects the modular yet interdependent nature of the models. Shared features enable synergistic learning across tasks, while preserving model-specific inputs which allows for task-specific optimization. This is particularly valuable in clinical settings were overlapping but non-identical decision goals must be simultaneously addressed (Figure 6).

**Figure 6:**
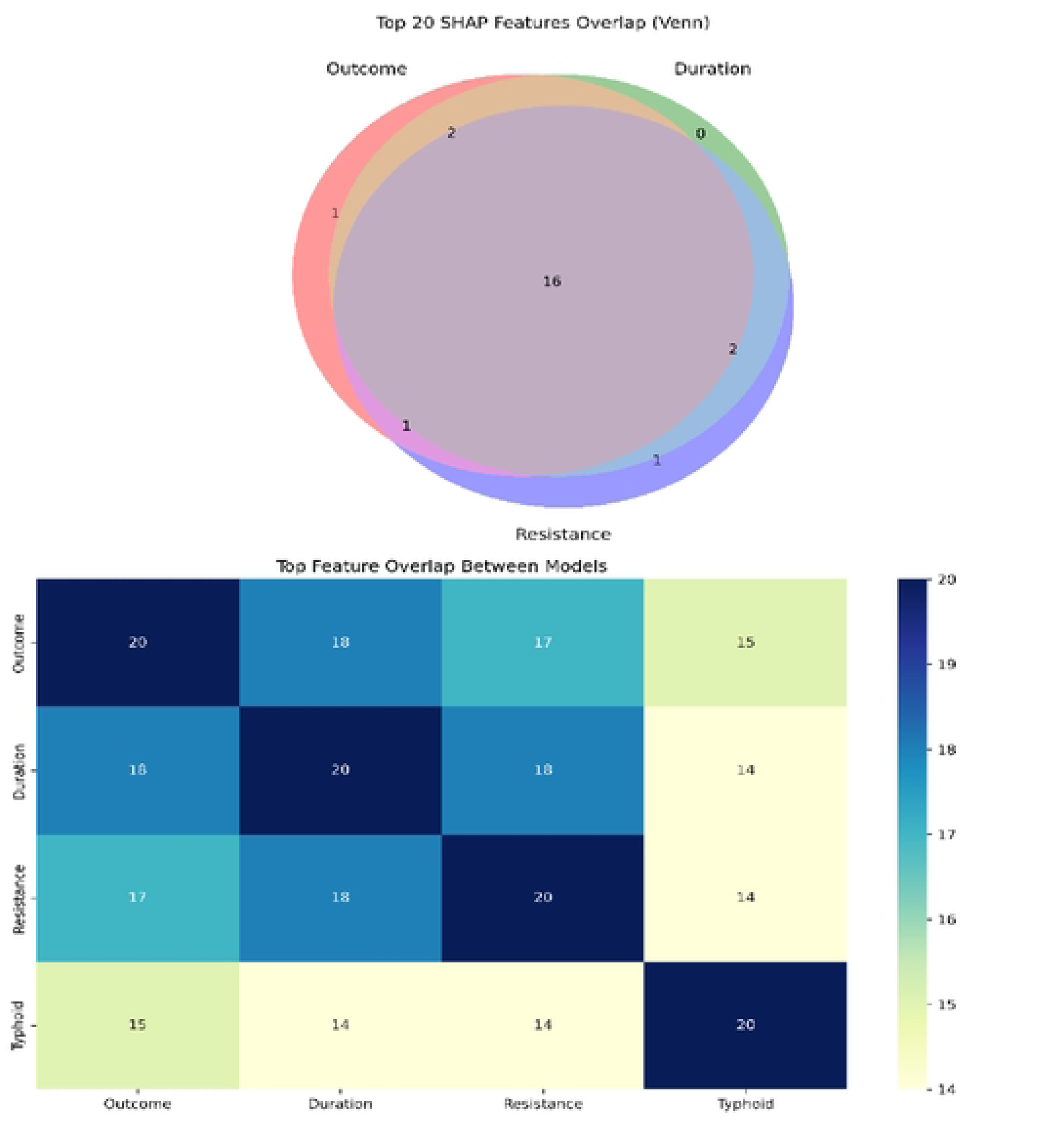
Cross-model SHAP feature, overlap across predictive tasks. Venn diagram (lop) and pairwise overlap heatmap (bottom) showing the number of shared top-ranked SHAP features across the four models. Common predictors such as Platelet Count, Age, PCI, and Hemoglobin(g/dL) are recurrent across models, while others remain task-specific. This pattern reflects a balance reflects shared physiological drivers and modular task-specific signals.

#### 3.6.3 Patient-Level Explanations and Personalized Attribution

To complement global feature importance analyses, patient-level explanations were generated using SHAP across all four predictive models. Each model was visualized through standardized layouts combining summary plots and individual waterfall plots (Figures S5–S8), which together illustrate how global feature contributions translate to personalized predictions. This methodology follows best practices for model-agnostic explanation generation and supports individualized insight into complex ensemble decisions (70).

For the Treatment Outcome model (Figure S5), the individualized attribution plot demonstrates that platelet count, potassium, and age positively contribute to the model results hence collectively shifted the probability toward recovery, reinforcing their relevance as interpretable clinical markers of prognosis. Similarly, the Treatment Duration model (Figure S6) patient-level decomposition revealed that platelet count, and hemoglobin are the main positive duration predictors but with nuanced contributions of age and biochemical measures in shaping the final estimate. The Resistance model (Figure S7) showed a greater reliance on the culture type in the negative direction and biochemical features, particularly calcium and potassium, alongside platelet count in the positive direction. These markers influenced the individualized resistance score in a manner consistent with reported associations between culture type, electrolyte imbalance and antimicrobial resistance phenotypes (24,27). Finally, for the Typhoid classification model (Figure S8), Patient 0 SHAP shows that SimMet_EnergyDisruption had a positive contribution to the model while severity score shows a negative contribution to model results, normalized duration and potassium also had negative contribution to the model in the patient-level waterfall plot providing a transparent account of how the model aggregated clinical and demographic signals into a binary decision.

The SHAP-based analysis confirms that interpretable modeling can be achieved in multi-task infectious disease prediction without compromising predictive power. model-specific emphasis, such as the Resistance model’s focus on Culture type versus the Typhoid classifier’s integration of broader severity-related variables, suggests that different predictive tasks leverage distinct, but clinically plausible, biological pathways. The strong influence of hematological (Platelet Count, Hemoglobin) and biochemical features (Calcium, Potassium) reflects their central role in patient stratification and treatment response estimation, corroborating findings from prior clinical studies on typhoid and sepsis outcomes while extending interpretability frameworks into the domain of personalized infectious disease prediction (73). The consistent prominence of platelet count across all models underscores its potential as a unifying biomarker for infection severity, treatment duration, and recovery trajectory.

#### 3.6.4 Attribution-Based Patient Clustering

By concatenating SHAP value vectors across all four models, we projected patient-level explanatory profiles into two dimensions using UMAP and t-SNE (Figure 7). Both embeddings revealed distinct, reproducible clusters representing latent patient subgroups defined by shared prediction logic rather than raw clinical inputs. For example, one cluster was enriched for patients with high severity indices, anemia, and positive resistance scores, suggestive of a high-risk metabolic/immunological subtype. The consistency of this signal across embedding techniques supports its robustness. This analysis demonstrates how explainability-informed clustering can uncover clinically meaningful substructures within heterogeneous infectious disease cohorts. Such an approach extends beyond model performance to enable AI-assisted triage, hypothesis generation, and the identification of explainability-guided patient subtypes, in line with emerging literature on interpretable machine learning for precision medicine (74).

**Figure 7:**
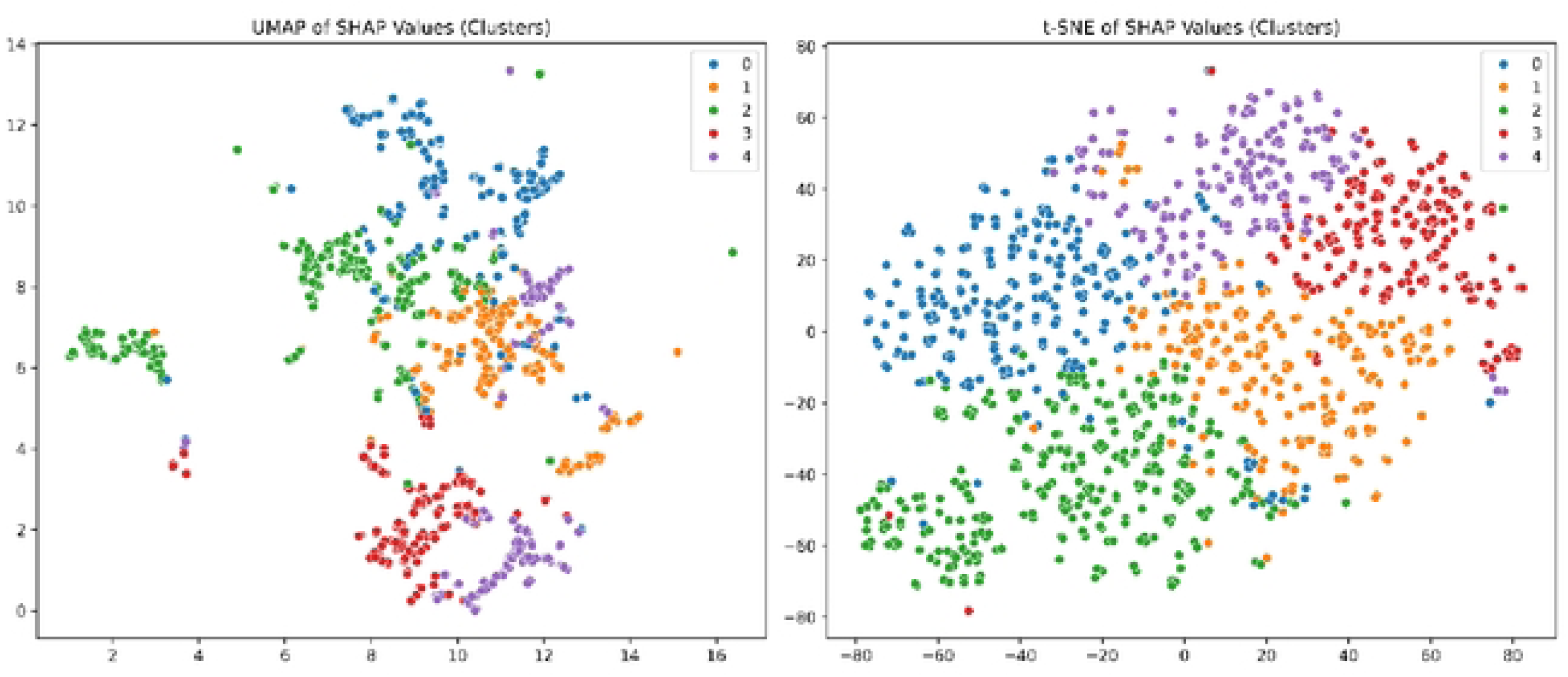
Low-dimensional projection of patient-level SHAP value vectors across all models using UMAP and t-SNE.

### 3.7 Drug Simulation Recommendations

#### 3.7.1 Model-Wide and Per-Drug SHAP Interpretability

Using the same underlying outcome model, we grouped patients by their recommended drug and used SHAP analysis to evaluate the model’s reasoning within each subgroup. Figures 8 and S9 show the results for both global and drug-specific explanations of prediction behavior after simulating treatment outcome probabilities for each patient across all candidate antibiotics. In line with previous clinical findings, they all displayed the same primary two features: platelet count and age, while the other three primary features were potassium, calcium, and hemoglobin, but in a different order (different SHAP magnitudes). Thus, the physiology of the patient played a significant role in all recommendations. This analysis reveals both shared and drug-specific patterns in treatment decision logic. Consistently across all groups, Platelet Count and Age were the most influential features.

**Figure 8:**
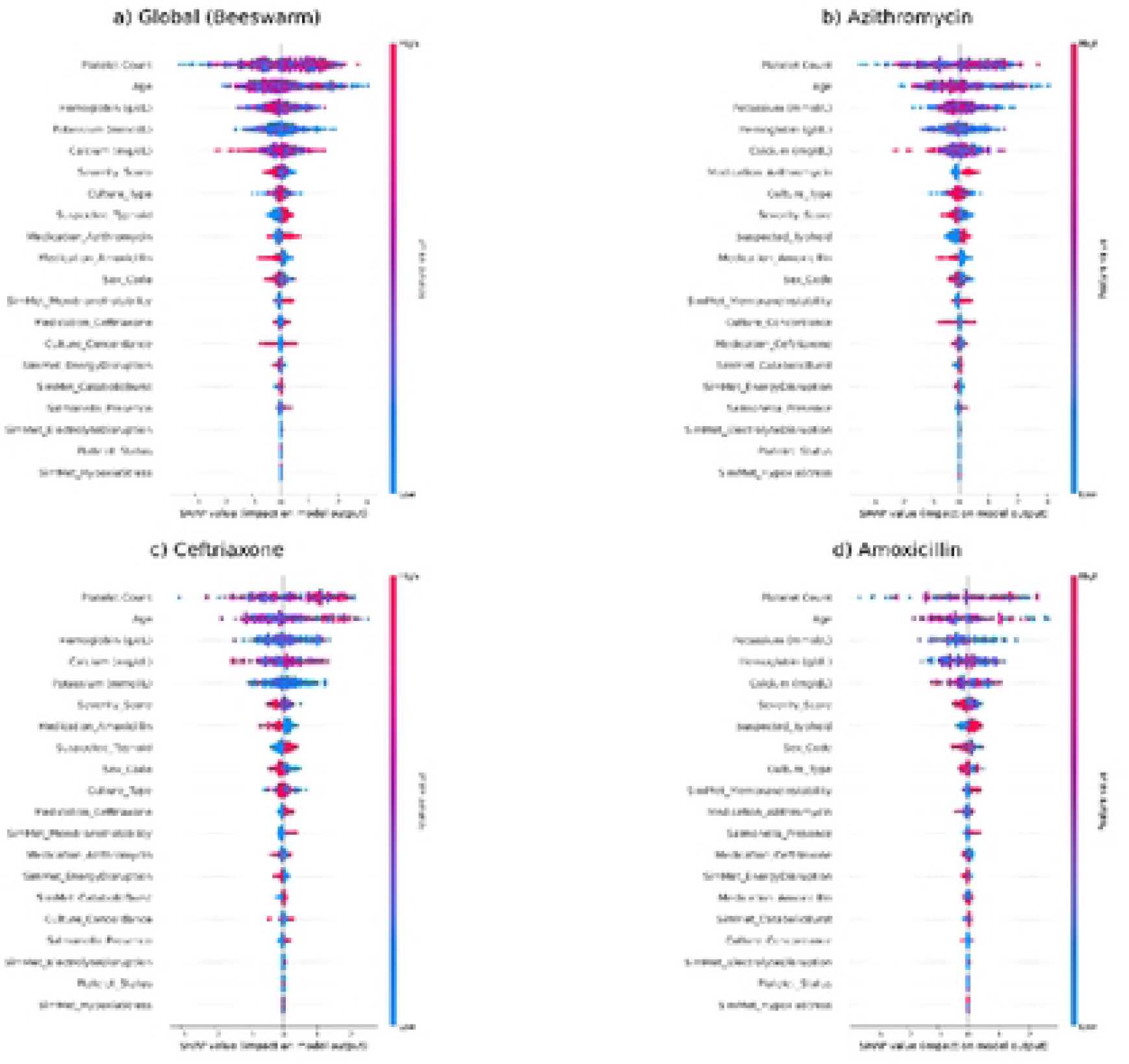
SHAP beeswarm plots showing feature contributions to treatment outcome predictions globally (a) and stratified by Top-1 predicted drug: Azithromycin (b), Ceftriaxone (c), and Amoxicillin (d).

Stratified analysis revealed subtle variances despite this overlap: recommendations for Ceftriaxone and Azithromycin were more impacted by diagnostic indicators, whilst Amoxicillin seemed to be more closely associated with stable physiological markers. It is interesting to note that the SHAP impact of culture outcomes and simulated metabolomic stress factors was consistently low across all groups. This implies that, under the current modeling settings, these variables offer small value. This could be because of data sparsity or redundancy with key clinical markers.

#### 3.7.2 Top vs Actual Drug

We compared the top predicted medications with observed treatment records in order to assess the degree of agreement between model-generated drug recommendations and real clinical prescriptions. The evaluation contrasted the actual drug/prescribed drug that clinicians prescribed with the Top Drug prediction (Top1 Match means the Top1_Drug matches the Actual_Drug, Top2 Match means the Top2_Drug matches the Actual_Drug, but Top1_Drug does not, and No Match means neither the Top1_Drug nor the Top2_Drug matches the Actual_Drug), which was the model’s main recommendation. Using a weighted average technique to account for class imbalance, we calculated important classification measures such as accuracy, precision, recall, and F1-score. Additionally, Seaborn’s heatmap tool was used to show the confusion matrices. This analysis captures not only exact Top-1 matches but also near-miss recommendations where the correct drug appeared as the model’s second-best suggestion, which is valuable in multi-option clinical decision-making contexts.

The final distribution of match types was summarized using absolute counts and normalized percentages to facilitate comparison across categories. To further evaluate how alignment between model-predicted drug recommendations and clinically administered drugs relates to treatment outcomes and key numeric indicators, and why the model may recommend a different drug where the actual outcome was successful, and find out whether it actually prioritized success probability, lower treatment duration, or resistance proxy score, we performed a stratified analysis based on match type. Subsequent analysis focused on comparing treatment success rates and quantitative prediction metrics across match types. Descriptive statistics (mean, standard deviation, and count) for these variables were computed per match type. A success rate was calculated as the proportion of patients in each group for whom the Treatment Outcome was labelled as successful. This analysis was designed to uncover systematic differences in predicted and real-world outcomes across model confidence levels, providing insight into the practical reliability of the Top-K drug recommender system. This type of comparative outcome analysis is commonly used in clinical decision support validation studies to assess model utility under partial match scenarios and helps contextualize predictive performance beyond standard classification metrics (75,76).

In 37.19% of cases, the model’s Top-1 predictions matched the actual medication that was provided, whereas in 27.24% of cases, the true medication was the model’s second-best recommendation. The top two predictions of the model did not match the recommended medication in 35.57% of cases. The Top-1 classification showed a weighted F1-score of 0.36 and an overall accuracy of 0.3719. Azithromycin performed quite well (recall: 0.57, precision: 0.38), Ceftriaxone performed moderately well (F1: 0.33), and Amoxicillin performed poorly (0.20). With no discernible performance benefit from using the second-ranked medication in the evaluation, the Top-2 classification produced somewhat lower performance metrics (accuracy: 0.2724, F1-score: 0.27; Tables S3–S4; Figures S10–S11).

Significant trends in treatment-related outcomes were found using a stratified match-type analysis. With an 89% treatment success rate, the Top-1 Match group was the most successful, followed by the Top-2 Match group (61%), and the No Match group (13%). Remarkably, the Top-2 Match group had the lowest predicted resistance proxy scores (mean: 8.55), indicating that the model occasionally gave antimicrobial stewardship precedence over strict adherence to prescription guidelines (Table S5; Figure 9). Model selection did not seem to be influenced by the predicted treatment durations, which were comparable across groups.

**Figure 9:**
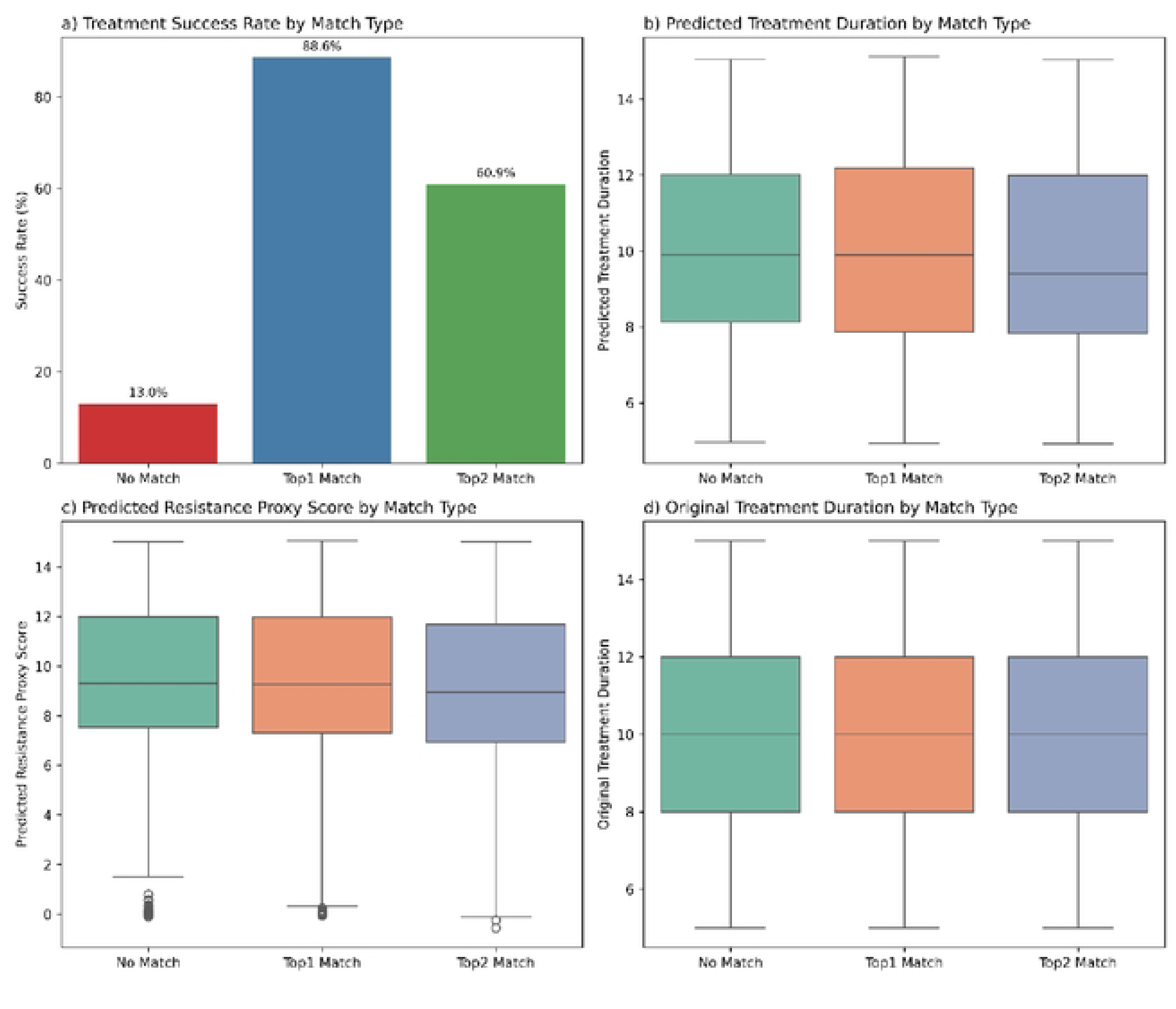
Summary of treatment outcomes and prediction characteristics by drug match type. Bar plots of treatment success rate a), predicted treatment duration b), predicted resistance score c), and original Treatment Duration across Top-1 Match, Top-2 Match, and No Match groups.

A crucial distinction in the interpretation of drug recommendation systems is brought to light by this evaluation: a departure from actual prescribing does not always signify model failure. Clinical recommendations are not always the best in terms of results, resistance reduction, or therapeutic effectiveness, even if they are frequently informed by experience and contextual information. Therefore, rather than misclassification, discrepancies between model and physician choices can be the result of informed prioritization. The highest success rate was recorded by patients in the Top-1 Match group, confirming that the model frequently corresponds with practical, efficacious prescriptions. Treatment effectiveness remained high (61%), and resistance proxy scores were lowest in the Top-2 Match group, although the genuine drug was not the model’s first recommendation. This suggests that the model may provide clinically viable alternatives with better resistance proxy profiles. This highlights the model’s potential to support antibiotic stewardship, a critical consideration in settings with rising antimicrobial resistance (42).

The No Match group, on the other hand, had worse treatment results and higher projected resistance ratings, highlighting the fact that complete disagreement between the model and doctors may frequently represent less-than-ideal circumstances. When it comes to model-aided decision review, this group might be particularly helpful in assisting clinicians in identifying situations where retrospective model insights point to the need for reconsideration. These findings are consistent with recent clinical AI research that highlights the importance of assessing decision-support tools based on their ability to improve health outcomes and system-level objectives like resistance control and duration reduction, in addition to classification accuracy (77). To fully realize this promise, outcome-aware optimization goals and ongoing feedback loops between model predictions and changing clinical standards must be included.

Analysis of initial treatment success across match types revealed clear and consistent patterns (Figure 10). According to preliminary data, 89.5% of patients (427 out of 482) in the Top1 Match group achieved treatment success when the model’s principal suggestion matched the actual medication administered. Success was noted in 60.9% of patients (215 out of 353) in the Top2 Match group. The No Match group, on the other hand, had a significantly lower success rate of 13.0% (60 out of 461). These findings imply that the model’s best predictions and successful clinical outcomes are highly aligned. The model’s potential as a helpful tool for assisting with treatment decisions is further highlighted by the significant drop in success rate that occurred when the top two recommendations from the model did not match the recommended medication. Crucially, the high success rate in the Top1 Match group (89.5%) offers compelling proof that results are typically positive when the model’s top recommendation coincides with clinical preference. The assumption that the model’s secondary recommendations would still be viable options that might be adjusted for additional variables like resistance profiles or wider generalizability is further supported by the Top2 Match group’s moderate success rate (60.9%).

**Figure 10:**
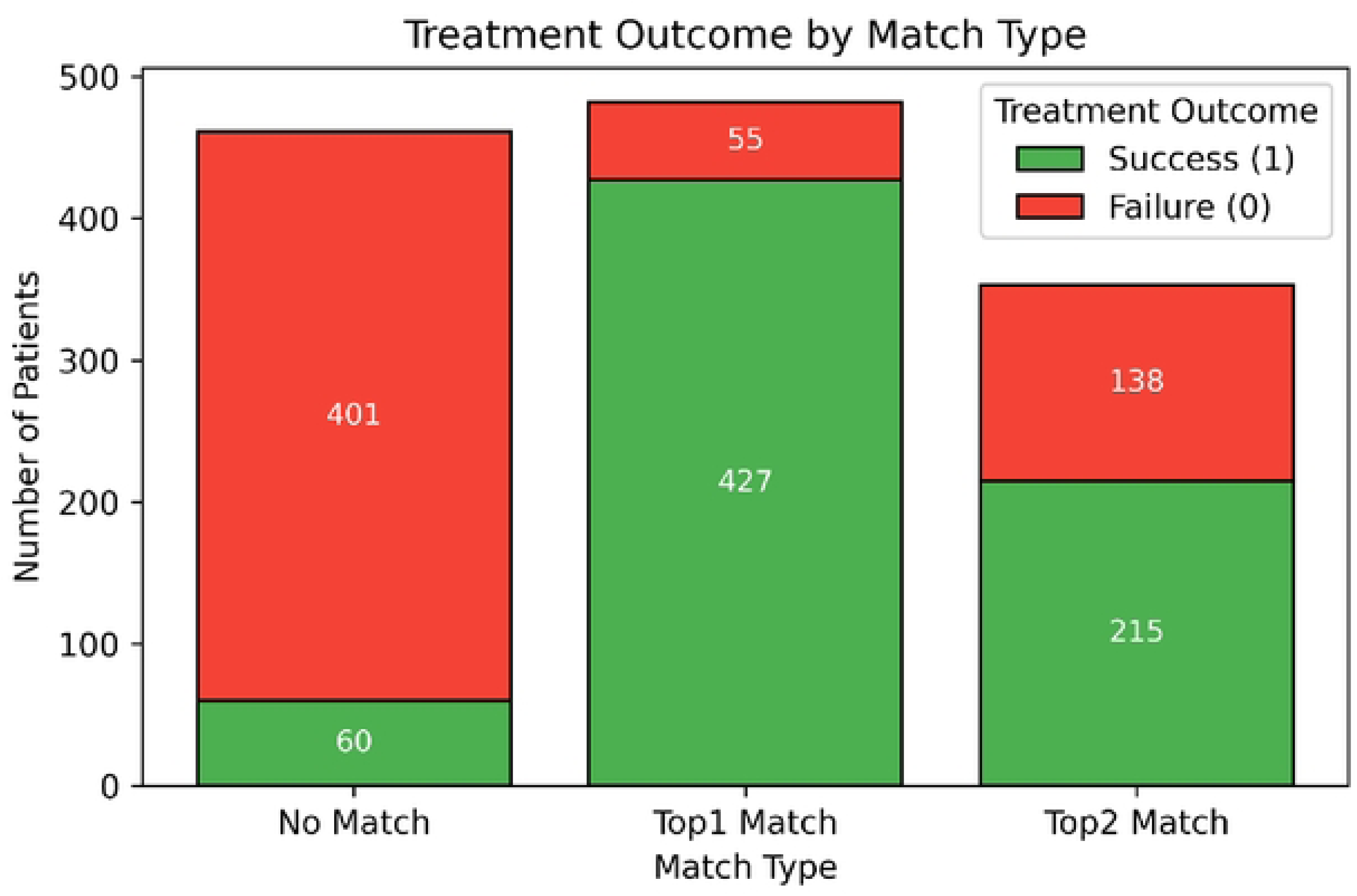
Treatment outcomes stratified by model-prescription match type. Stacked bar plot showing the number of initially successful and failed treatments within each match type group (Topi Match, Top2 Match, No Match). The highest success rate was observed in the Topi Match group (427/482), and the lowest in the No Match group (60/461).

On the other hand, significant concerns are raised by the low success rate of 13.0% in the No Match group. This may suggest instances when the model was more in line with outcome-driven trends in the data, but it does not always imply that the doctor made a mistake. Retrospective evaluation may be warranted in certain cases, particularly when resistance-driven failures or substantial clinical ambiguity are involved. Importantly, this analysis confirms that match alignment by itself is not a perfect standard; rather, the observed results support the model’s ability to suggest medications linked to better treatment outcomes. These results are in line with the growing belief that model evaluation in clinical decision support should take into account the downstream influence on health outcomes in addition to agreement with past decisions (78).

#### 3.7.3 Confidence analysis

We conducted two confidence quantification assessments to evaluate the certainty of AI-generated drug recommendations: Absolute Confidence Classification, which assessed the model’s anticipated success probability of the top-ranked drug (Top1_Probability) for each patient. The advice was deemed Confident if this value was ≥ 0.5 (see calibration) or Ambiguous otherwise. The calibration above demonstrated that predictions at 0.4 were close to the calibration standard, and this threshold is consistent with the conventional practice of treating outputs above 0.5 as high-certainty in probabilistic binary classification (79) (Table S6).

For Relative Confidence, we determined the confidence margin between the top two drug probabilities in order to evaluate model preference between medications that were closely ranked: Top1_Probability – Top2_Probability = Confidence Margin. In order to capture even little ranking dominance while being robust against prediction noise, a forecast was deemed confident if this margin was ≥ 0.01.

There is a clear bimodal pattern in the Top1_Probability histogram (Figure 11a). One distinct cluster of predictions is close to 1.0, which denotes great certainty, and another is close to zero. According to bar plots, 561 patients were classified as uncertain, while 735 out of 1296 patients obtained confident medicine recommendations based on Top1 projected probabilities (≥ 0.5) (Figure 11C).

**Figure 11:**
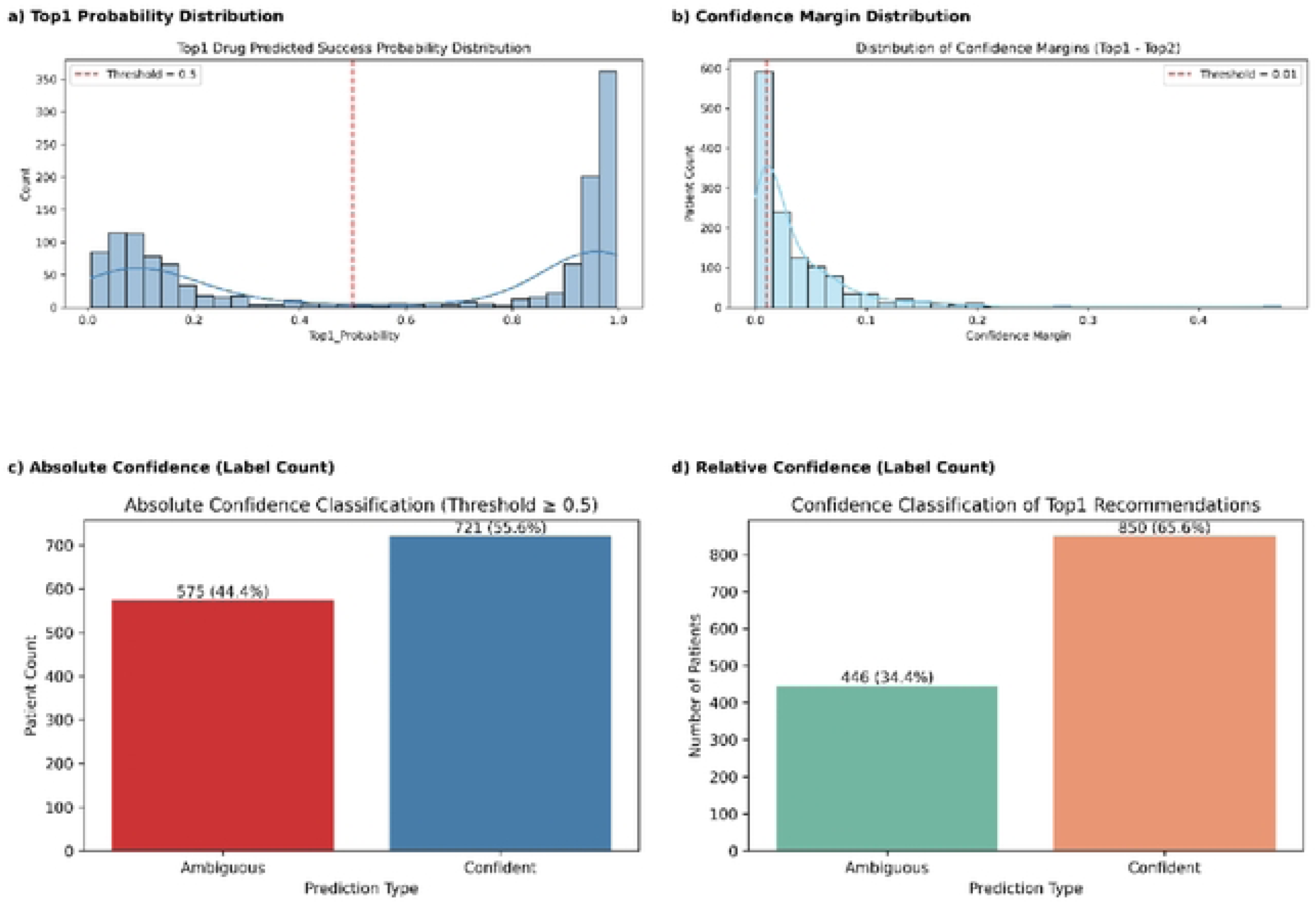
Confidence analysis of Topi drug recommendations across 1,296 patients. The predicted success probabilities (a) show a bimodal distribution, with a reference threshold of 0.4 used to illustrate model certainty levels. The confidence margin distribution (b), defined as the difference between the Topi and Top2 predicted probabilities, reveals a strong skew toward low margins, with a threshold of 0.01 applied to classify relative confidence. Subplots (c) and (d) summarize the absolute (Topl_Probability >0.5) and relative (margin > 0.01) confidence classifications, respectively, illustrating the proportion of predictions considered confident versus ambiguous under each criterion.

With a strong tail skewed towards smaller margins, most Top1–Top2 margins fall below 0.05. To strike a balance between inclusivity and model confidence, a criterion of 0.01 was applied (Figure 11b). This adjustment significantly raised the percentage of confident classifications from 48.5% (at 0.02) to 65.6%. 850 patients (65.6%) obtained confident predictions using the margin-based technique, demonstrating the advantage of employing a more sensitive threshold (0.01) for ranking clarity (Figure 11D).

#### 3.7.4 Treatment Discrepancy Analysis

We conducted a treatment discrepancy analysis, which simulates and compares the expected success probabilities between (i) the model’s Top1 drug recommendation and (ii) the medication that the clinician actually prescribed, in order to assess the dependability and potential clinical utility of model-driven treatment recommendations.

Using the prediction output, we first looked for instances when the model’s Top1 recommendation deviated from the observed clinical prescription. From a held-out test set, which included encoded drug vectors for simulation, patient-level clinical and biochemical data were acquired. By re-injecting the patient record and switching the drug vector to activate the clinician-selected medication (1 for the actual drug, 0 for others), we were able to mimic the success probability of the actual prescribed drug using an outcome model. We were able to calculate ΔProbability = P (Model Top1) – P (Actual Prescribed) as a result. A positive Δ indicates that, according to the internal patterns of the model, the drug suggested by the model has a higher chance of producing a successful course of therapy than the medication chosen by the doctor. Age, sex, inpatient duration, comorbidities, and pathogen-type metadata were added to a case-study subset of high-confidence differences (Δ > 0.05) to enhance interpretability and possible therapeutic translation.

In 814 (62.8%) of the 1,296 test patients, the model’s Top1 suggestion deviated from the medication provided by the doctor, resulting in inconsistent treatment choices. 100% (814/814) of all discrepant cases produced a positive Δ Probability, meaning that the model consistently projected better success likelihoods for the drugs it advised than for the prescription written by the doctor. The distribution of Δ values, which had a strong right-skew and varied from 0.001 to 0.67, is shown in Figure 12. A non-negligible tail went beyond 0.2, indicating extremely different forecasts for some individuals, whereas over 60% of Δ values were below 0.1. Ten high-confidence case studies (Δ > 0.05) were selected for qualitative review (Table S7). The model may be more sensitive to such high-risk configurations when choosing the best treatments because these patients tended to have multiple symptoms, underlying chronic diseases, or infections linked to high antibiotic resistance.

**Figure 12:**
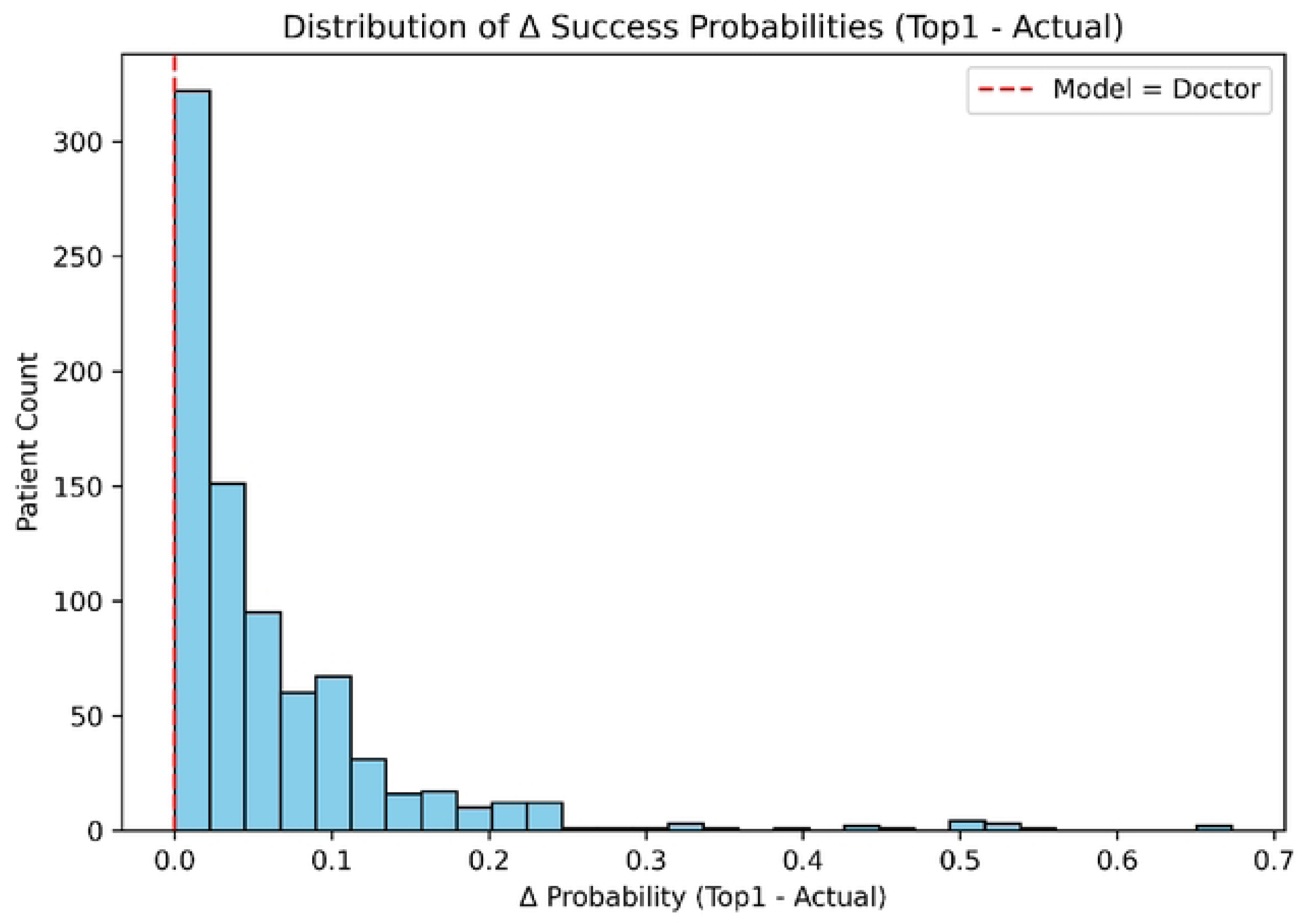
Distribution of Δ Success Probabilities (Topi – Actual). Histogram illustrating the distribution of predicted success probability differences between the model’s Topi drug recommendation and the clinician’s prescribed drug. A positive Δ value indicates higher predicted success for the model’s choice. The red dashed line (Δ = 0) represents cases where the model and clinician had equivalent predicted efficacy.

When there is disagreement, the machine learning model regularly suggests therapies that are expected to function better than the clinician’s selection, at least according to success probability derived from training data, as this discrepancy study shows. This suggests a possible discrepancy between data-driven optimization and empirical decision-making, but it does not imply clinical mistake. The potential of AI-guided medication selection in pharmacogenomics, cancer, and infectious illnesses has been highlighted by a number of research (77,78,80,81). Integrating probabilistic models provides a potent way to assist clinicians in the context of infectious disease, where patient variability and resistance patterns can significantly change outcomes, especially in environments with high levels of uncertainty or inadequate antimicrobial stewardship infrastructure.

A clinically interpretable proxy for confidence in switching to the model’s advice is the Δ Probability metric. The model’s calibration is suitable for all drug classes, and the encoded patient-drug interaction accurately depicts biological dynamics, although its dependability depends on a number of assumptions. Unmeasured confounding, such as doctor intuition or contextual knowledge that is not recorded in features, does not exist. Therefore, before any therapeutic integration, prospective validation in real-world situations is essential, even though the model was statistically favored in 100% of discrepant cases. Future research should include mixed-methods prescriber feedback, causal inference frameworks, and uncertainty quantification.

#### 3.7.5 Risk Triage

We performed a stratified risk triage analysis using antibiotic resistance proxy scores and estimated treatment time derived from a machine learning method. Four different strata, designated Q1 (Shortest) through Q4 (Longest), were created when the duration forecasts were divided into quartiles. To examine trends in risk escalation, we computed average resistance proxy scores, patient numbers, mean, minimum, and maximum treatment durations per quartile. The triage framework’s equal categorization of patients across quartiles led to a monotonic increase in both expected duration and resistance risk (n = 324 per group). Figure 13 and Table S8 show a direct correlation between longer treatment durations and higher resistance proxy scores, suggesting a co-progression of resistance risk and therapeutic demand.

**Figure 13:**
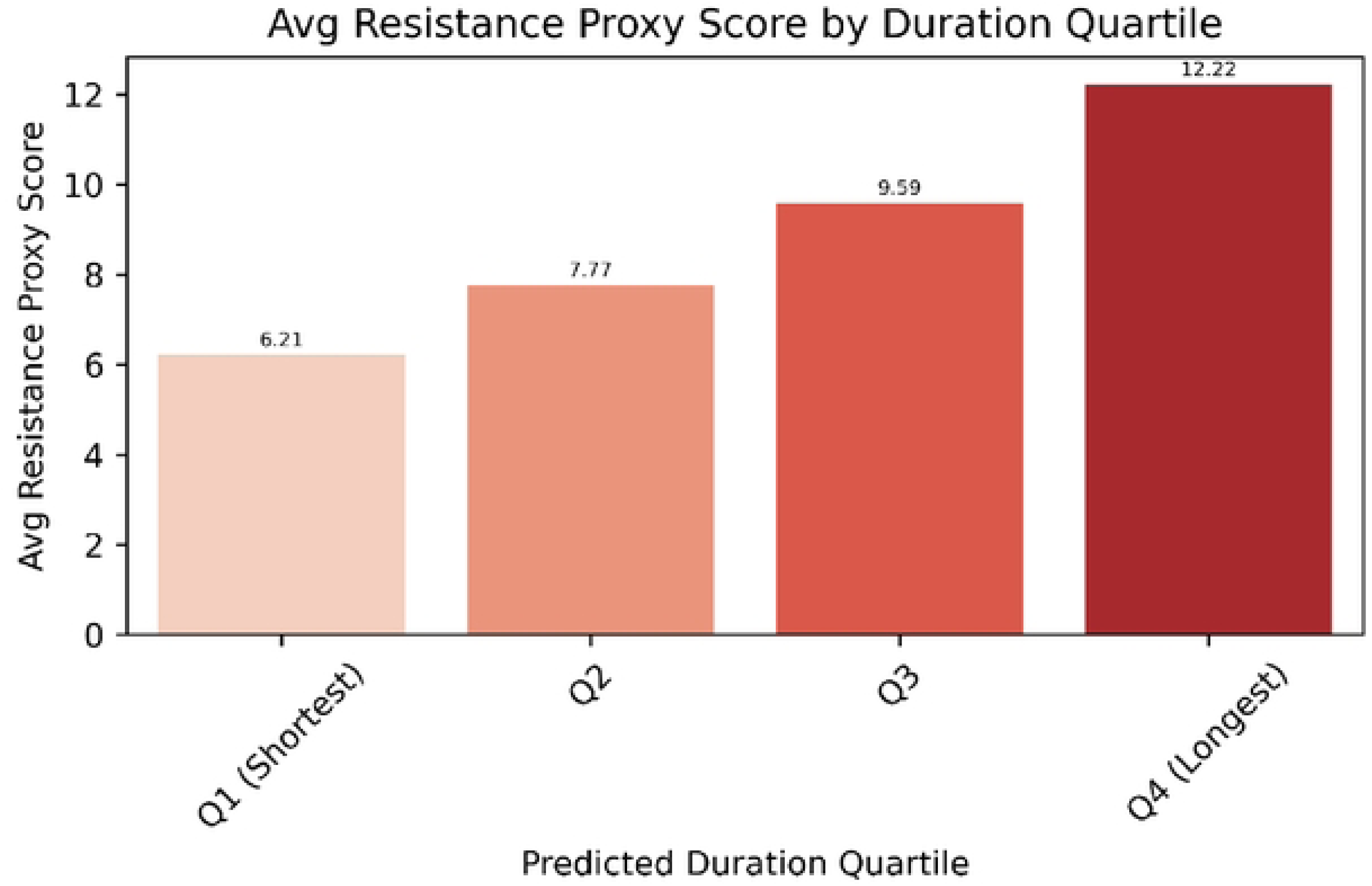
Mean resistance proxy scores stratified by predicted treatment duration quartiles (Ql-Q4). Quartiles are labeled based on ascending treatment duration estimates. A stead) increase in resistance score across quartiles is observed, indicating risk co-escalation.

A practical method for identifying patients at increasing risk based on resistance likelihood and length burden is offered by stratified triage. For example, patients in Q4 have longer expected durations (mean: 13.41 days) and higher resistance scores (mean: 12.22), which make them crucial profiles that need greater monitoring and maybe second-line medicines. This is consistent with previous research showing that resistant or recurrent infections are frequently linked to extended therapy (5,82,83). Clinicians can reveal feature-level causes for prolonged durations by utilizing SHAP-based interpretability, which promotes openness and focused intervention planning. A patient who was recognized in Q4 for having underlying chronic diseases and high severity, for instance, may be preemptively prioritized for aggressive therapy modification or adjunct diagnostics. This method enables proactive patient segmentation, guiding antimicrobial stewardship, especially in low-resource settings where empirical decision-making often lacks algorithmic support.

#### 3.7.6 Per Drug summary analysis

Using predictions from our treatment outcome model, we conducted a per-drug stratified study to evaluate drug-specific performance and model interpretation. For summary data, such as mean success rate, expected treatment duration, resistance proxy score, and Top1 likelihood (model confidence), patients were categorized according to the medications they actually received. Amoxicillin, azithromycin, and ceftriaxone had counts of 437, 465, and 394, respectively (Figure 14 and Table S9). Each medicine had a different mean Top1 probability and treatment success rate. With the greatest average model confidence (Top1 = 0.603) and the highest success rate (58.5%), azithromycin was the best-performing medication. Ceftriaxone had the lowest clinical success rate and model confidence.

**Figure 14:**
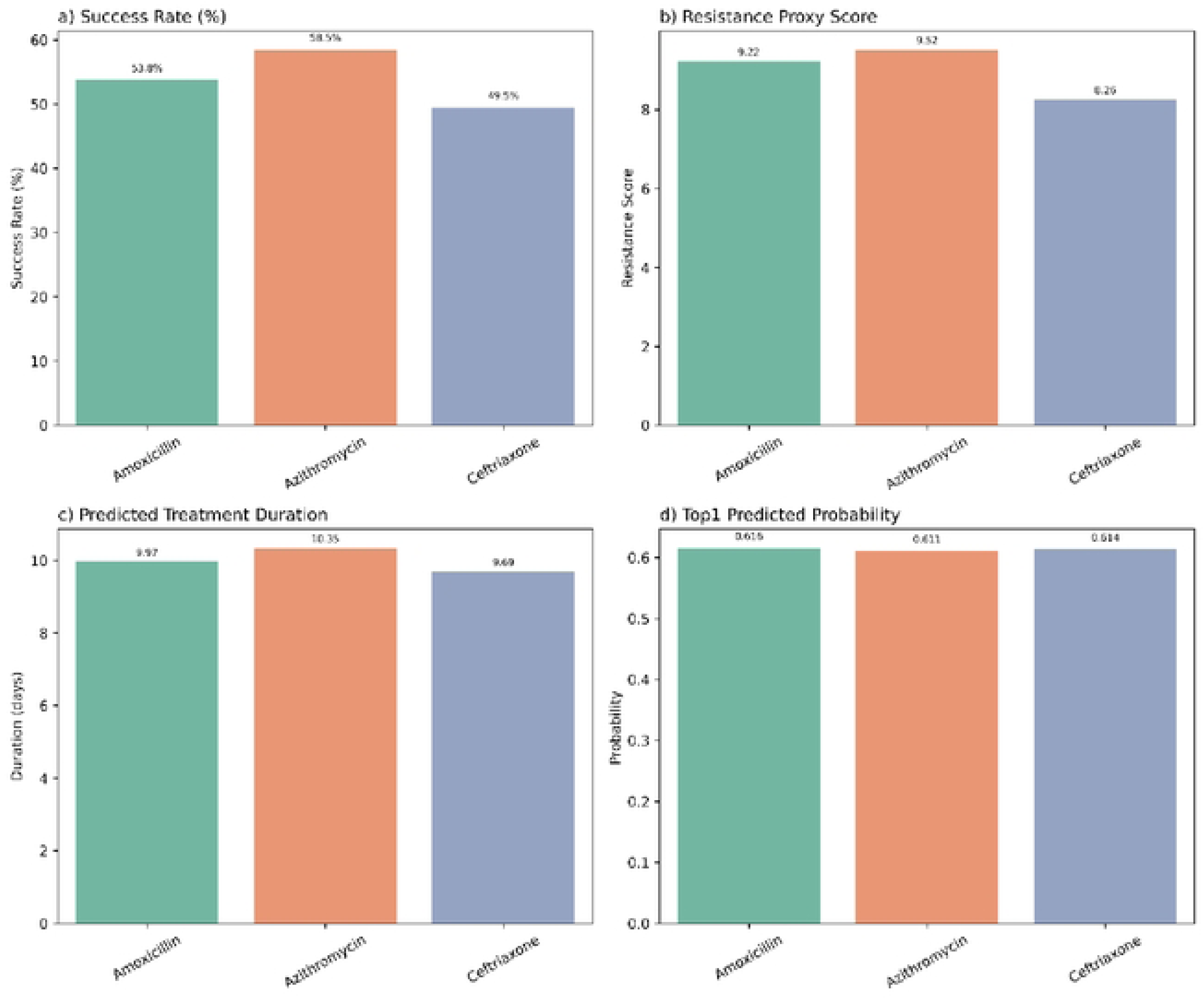
presents a comparative grid of four key indicators: success rate, resistance proxy score, predicted duration, and Topi probability per drag. These differences underscore potential optimization opportunities in antibiotic stewardship.

This analysis shows how SHAP and outcome stratification can be used to bridge the gap between model interpretability and clinical relevance. Antimicrobial resistance trends reported in typhoid treatment studies are mirrored by the higher resistance proxies and superior predicted success rates of drugs such as azithromycin, which may be due to selective pressure or previous exposure patterns (72,84). As a quantitative stand-in for dependability, model confidence (Top1 probability) can be used to pinpoint treatments with unclear forecasts. Individualized feature breakdowns made possible by SHAP integration can be utilized to improve antimicrobial policy and clinical decision support systems.

We repeated the simulation process using the resistance and duration models (minimum predicted score/value) in addition to the outcome model in order to further refine our simulation predictions. When the results were compared to those of the outcome model alone, it was found that they were identical.

##### 4 Limitations and Future Directions

Despite promising performance and interpretability, several limitations must temper the conclusions of this study. First, although synthetic data were generated using nearest-neighbour sampling to preserve marginal distributions, joint feature distributions may still be imperfectly represented. This is particularly relevant for rare clinical constellations such as co-occurring severe presentations, specific antibiotic exposures, and treatment failure where even small deviations in joint structure can influence model behavior. Binary thresholding of clinical variables, while pragmatic, can introduce discontinuities near decision boundaries; small fluctuations around these cut-points may produce unstable classification behavior. Similarly, the PCA-derived components used for dimensionality reduction may obscure biologically meaningful correlations even as they capture dominant covariance structure. Future work should explore context-aware generative models such as conditional tabular GANs (CTGANs) or variational autoencoders conditioned on clinical subgroups to better preserve higher-order dependencies. Integrating supervised or pathway-informed dimensionality reduction approaches may also help align latent structure with biologically interpretable processes.

Model uncertainty and calibration constitute a second constraint. We found systematic overestimation in high-confidence locations, despite the overall calibration being satisfactory. Previous research demonstrates that post-hoc calibration techniques like isotonic regression or Platt scaling significantly increase the probabilistic reliability of boosted tree models, including XGBoost. These techniques should be included in subsequent rounds. Furthermore, our models may not adequately account for unmeasured confounders because they only use observed factors such symptoms, vital signs, laboratory markers, and demographics. Comorbidities, pharmacokinetic fluctuation, prescriber judgment, and treatment adherence are examples of factors that may concurrently affect therapy choice and results but were not recorded. Although the simulation framework provides a useful way to examine counterfactual drug allocations, it implicitly assumes conditional exchangeability: that treatment groups can be compared fairly after adjusting for observed features. This assumption is unlikely to hold completely in routine clinical settings, and our ΔProbability estimates should therefore be interpreted as *associational*, not causal. Future work should explore formal causal inference frameworks, including directed acyclic graph (DAG)-guided adjustment, propensity score or inverse probability weighting, and where available, laboratory gold standards such as AST or MIC data to improve causal validity (85,86).

Generalizability is a third drawback. The analysis does not include external or temporal validation and is based on a single-center cohort. Models trained in one context may perform poorly when used in another due to the dynamic nature of antibiotic resistance and changing clinical practice. Temporal dependencies that our cross-sectional approach is unable to capture would be better captured by longitudinal modeling of illness trajectories. Prior to any clinical implementation, model updating, recurring recalibration, and data drift observation will be crucial. Lastly, no ML system should be regarded as prescriptive without thorough future validation, even though our simulation findings appear promising. The necessity for professional infectious disease oversight is highlighted by the unpredictable failure of clinical AI systems in unusual or under sampled cases. Safe implementation will require institutional readiness, including ethical governance, stakeholder consultation, regulatory compliance, transparent reporting, and mechanisms to ensure equity and patient privacy (87).

##### 5. Conclusion

This study demonstrates that machine-learning models can meaningfully support antibiotic selection for typhoid fever by leveraging routinely collected clinical data and patient-level phenotypes. Across multiple predictive tasks, the framework identified individualized treatment strategies that were frequently associated with higher predicted success probabilities than the clinician-prescribed regimens observed in the dataset. These findings do not imply clinical error; rather, they illustrate how data-driven optimization can complement empirical prescribing and highlight situations where alternative therapies might merit consideration. The ΔProbability measure, in particular, offers a practical way to visualize potential improvements from drug switching, although its interpretation must remain associational until validated prospectively.

The framework underscores the potential of AI-guided prescribing to enhance antimicrobial stewardship in settings where resources are constrained and patient presentations are heterogeneous. Nonetheless, the system remains exploratory. Translation to clinical practice will require external validation across diverse epidemiological settings, integration with microbiological and genomic resistance data, and mixed-methods engagement with frontline clinicians to ensure interpretability, usability, and safety. With these steps, machine-learning approaches could evolve into reliable decision-support tools that augment but not replace clinical expertise in the management of infectious diseases.

##### Author contributions

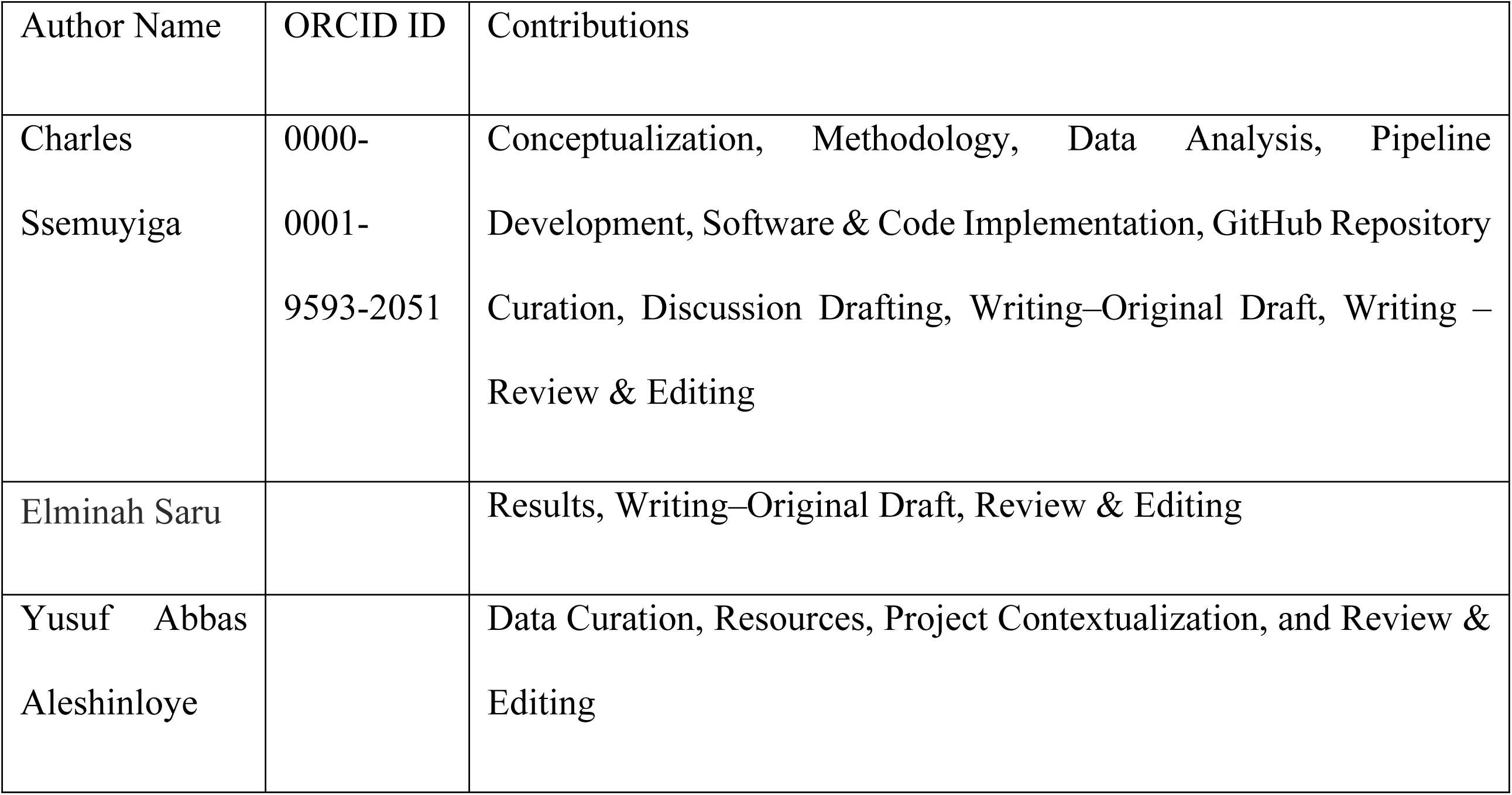

##### Conflict of interest

The author declares no commercial or financial relationships that could be construed as a potential conflict of interest.

##### Data availability statement

Publicly available datasets were analyzed in this study. This data can be found here: https://github.com/SsemuyigaMHC/TyphoidRx

##### Funding

This research received no specific grant from any funding agency in the public, commercial, or not-for-profit sectors.

##### Supplementary Material

Supplementary figures and tables supporting this article are provided in the online supplementary file. These include detailed distributions of Δ Probability values, case study subsets, and extended methodological notes.

## Acknowledgments

The authors thank the PharmaQsar Bioinformatics Firm for support and resources provided during the project.

## Generative AI Statement

The authors declare that no generative artificial intelligence (AI) was used in the creation of this manuscript. Paraphrasing software (QuillBot) was employed exclusively for stylistic editing and did not contribute to content generation, interpretation, or analysis.

## References

1. Buzilă ER, Dorneanu OS, Trofin F, Sima CM, Iancu LS. Assessing Salmonella Typhi Pathogenicity and Prevention: The Crucial Role of Vaccination in Combating Typhoid Fever. International Journal of Molecular Sciences 2025, Vol 26, Page 3981 [Internet]. 2025 Apr 23 [cited 2025 Sep 7];26(9):3981. Available from: https://www.mdpi.com/1422-0067/26/9/3981/htm

2. Boakye Okyere P, Twumasi-Ankrah S, Newton S, Nkansah Darko S, Owusu Ansah M, Darko E, et al. Risk Factors for Typhoid Fever: Systematic Review. JMIR Public Health Surveill 2025;11:e67544 https://publichealth.jmir.org/2025/1/e67544 [Internet]. 2025 Aug 28 [cited 2025 Sep 7];11(1):e67544. Available from: https://publichealth.jmir.org/2025/1/e67544

3. Parry CM, Hien TT, Dougan G, White NJ, Farrar JJ. Typhoid fever. N Engl J Med [Internet]. 2002 Nov 28 [cited 2025 May 6];347(22):1770–82. Available from: https://pubmed.ncbi.nlm.nih.gov/12456854/

4. Crump JA, Mintz ED. Global trends in typhoid and paratyphoid fever. Clinical Infectious Diseases [Internet]. 2010 Jan 15 [cited 2025 May 6];50(2):241–6. Available from: https://pubmed.ncbi.nlm.nih.gov/20014951/

5. Ssemuyiga C. In-vitro determination of antibacterial activity of Cannabis Sativa against staphylococcus aureusIn-vitro determination of antibacterial activity of Cannabis Sativa against staphylococcus aureus. 2019 [cited 2024 Jan 21]; Available from: http://dissertations.mak.ac.ug/handle/20.500.12281/7027

6. Zhang X, Zhang D, Zhang X, Zhang X. Artificial intelligence applications in the diagnosis and treatment of bacterial infections. Front Microbiol [Internet]. 2024 [cited 2025 Sep 9];15:1449844. Available from: https://pmc.ncbi.nlm.nih.gov/articles/PMC11334354/

7. Charles S, Mahapatra RK. Artificial intelligence based de-novo design for novel Plasmodium falciparum plasmepsin (PM) X inhibitors. J Biomol Struct Dyn [Internet]. 2023 Nov 9 [cited 2023 Nov 9];1–16. Available from: https://www.tandfonline.com/doi/abs/10.1080/07391102.2023.2279700

8. Awe OI, Obura H, Ssemuyiga C, Mudibo E, Mwanga MJ. Enhanced Deep Convolutional Neural Network for SARS-CoV-2 Variants Classification. Front Artif Intell. 8:1512003.

9. Klemm EJ, Shakoor S, Page AJ, Qamar FN, Judge K, Saeed DK, et al. Emergence of an Extensively Drug-Resistant Salmonella enterica Serovar Typhi Clone Harboring a Promiscuous Plasmid Encoding Resistance to Fluoroquinolones. journals.asm.orgEJ Klemm, S Shakoor, AJ Page, FN Qamar, K Judge, DK Saeed, VK Wong, TJ DallmanMBio, 2018•journals.asm.org [Internet]. 2018 Jan 1 [cited 2025 Sep 9];9(1). Available from: https://journals.asm.org/doi/abs/10.1128/mbio.00105-18

10. Pennisi F, Pinto A, Ricciardi GE, Signorelli C, Gianfredi V. The Role of Artificial Intelligence and Machine Learning Models in Antimicrobial Stewardship in Public Health: A Narrative Review. Antibiotics 2025, Vol 14, Page 134 [Internet]. 2025 Jan 30 [cited 2025 Sep 9];14(2):134. Available from: https://www.mdpi.com/2079-6382/14/2/134/htm

11. Alami H, Lehoux P, Denis JL, Motulsky A, Petitgand C, Savoldelli M, et al. Organizational readiness for artificial intelligence in health care: insights for decision-making and practice. J Health Organ Manag [Internet]. 2021 Feb 5 [cited 2025 Sep 10];35(1):106–14. Available from: 10.1108/JHOM-03-2020-0074

12. de la Lastra JMP, Wardell SJT, Pal T, de la Fuente-Nunez C, Pletzer D. From Data to Decisions: Leveraging Artificial Intelligence and Machine Learning in Combating Antimicrobial Resistance – a Comprehensive Review. J Med Syst [Internet]. 2024 Dec 1 [cited 2025 Sep 9];48(1):1–14. Available from: https://link.springer.com/article/10.1007/s10916-024-02089-5

13. Algain S, Marra AR, Kobayashi T, Marra PS, Celeghini PD, Hsieh MK, et al. Can we rely on artificial intelligence to guide antimicrobial therapy? A systematic literature review. Antimicrobial Stewardship & Healthcare Epidemiology [Internet]. 2025 Mar 31 [cited 2025 Sep 9];5(1):e90. Available from: https://www.cambridge.org/core/journals/antimicrobial-stewardship-and-healthcare-epidemiology/article/can-we-rely-on-artificial-intelligence-to-guide-antimicrobial-therapy-a-systematic-literature-review/8239BEF5A37E8747203593A2D6C99DAE

14. Maviglia R, Michi T, Passaro D, Raggi V, Bocci MG, Piervincenzi E, et al. Machine Learning and Antibiotic Management. Antibiotics [Internet]. 2022 Mar 1 [cited 2025 Sep 10];11(3):304. Available from: https://www.mdpi.com/2079-6382/11/3/304/htm

15. Ribers MA, Ullrich H, Arpi RM, Bjerrum L, Cristina G, Currea C, et al. Battling Antibiotic Resistance: Can Machine Learning Improve Prescribing? SSRN Electronic Journal [Internet]. 2019 Jun 5 [cited 2025 Sep 10]; Available from: https://arxiv.org/pdf/1906.03044

16. Revell AD, Wang D, Wood R, Morrow C, Tempelman H, Hamers RL, et al. An update to the HIV-TRePS system: the development and evaluation of new global and local computational models to predict HIV treatment outcomes, with or without a genotype. Journal of Antimicrobial Chemotherapy [Internet]. 2016 Oct 1 [cited 2025 Sep 10];71(10):2928. Available from: https://pmc.ncbi.nlm.nih.gov/articles/PMC5031919/

17. Chen T, Guestrin C. XGBoost: A scalable tree boosting system. Proceedings of the ACM SIGKDD International Conference on Knowledge Discovery and Data Mining [Internet]. 2016 Aug 13 [cited 2025 May 20];13-17-August-2016:785–94. Available from: https://dl.acm.org/doi/pdf/10.1145/2939672.2939785

18. Lundberg SM, Erion G, Chen H, Degrave A, Prutkin JM, Nair B, et al. From local explanations to global understanding with explainable AI for trees. nature.comSM Lundberg, G Erion, H Chen, A DeGrave, JM Prutkin, B Nair, R Katz, J HimmelfarbNature machine intelligence, 2020•nature.com [Internet]. 2020 [cited 2025 Sep 10];2(1):56–67. Available from: https://www.nature.com/articles/s42256-019-0138-9.

19. Rajkomar A, Dean J, Medicine IKEJ of, 2019 undefined. Machine learning in medicine. Mass Medical SocA Rajkomar, J Dean, I KohaneNew England Journal of Medicine, 2019•Mass Medical Soc [Internet]. 2019 Apr 4 [cited 2025 Sep 10];380(14):1347–58. Available from: https://www.nejm.org/doi/abs/10.1056/NEJMra1814259

20. Shickel B, Tighe P, … ABI journal of, 2017 undefined. Deep EHR: a survey of recent advances in deep learning techniques for electronic health record (EHR) analysis. ieeexplore.ieee.orgB Shickel, PJ Tighe, A Bihorac, P RashidiIEEE journal of biomedical and health informatics, 2017•ieeexplore.ieee.org [Internet]. [cited 2025 Sep 10]; Available from: https://ieeexplore.ieee.org/abstract/document/8086133/

21. Carson JL, Grossman BJ, Kleinman S, Tinmouth AT, Marques MB, Fung MK, et al. Red blood cell transfusion: A Clinical practice guideline from the AABB. Ann Intern Med [Internet]. 2012 [cited 2025 May 3];157(1):49–58. Available from: https://pubmed.ncbi.nlm.nih.gov/22751760/

22. Platelet Count (PLT): Normal Range, Test Results & Meaning [Internet]. [cited 2025 May 3]. Available from: https://my.clevelandclinic.org/health/diagnostics/21782-platelet-count

23. Sadiq NM, Anastasopoulou C, Patel G, Badireddy M. Hypercalcemia. StatPearls [Internet]. 2024 May 7 [cited 2025 May 9]; Available from: https://www.ncbi.nlm.nih.gov/books/NBK430714/

24. Schafer AL, Shoback DM. Hypocalcemia: Diagnosis and Treatment. Endotext [Internet]. 2016 Jan 3 [cited 2025 May 9]; Available from: https://www.ncbi.nlm.nih.gov/books/NBK279022/

25. Calcium Blood Test: What It Is & Results [Internet]. [cited 2025 May 3]. Available from: https://my.clevelandclinic.org/health/diagnostics/22021-calcium-blood-test

26. Rastegar A. Serum Potassium. BMJ [Internet]. 1990 Dec 1 [cited 2025 May 9];2(4743):1341–2. Available from: https://www.ncbi.nlm.nih.gov/books/NBK307/

27. Potassium Levels Blood Test: High vs. Low, Normal K Level [Internet]. [cited 2025 May 3]. Available from: https://www.webmd.com/a-to-z-guides/potassium-blood-test

28. Typhoid Fever-Symptoms, Treatment, Causes and Diagnosis [Internet]. [cited 2025 May 9]. Available from: https://www.careinsurance.com/blog/health-insurance-articles/typhoid-how-dangerous-it-could-be?utm_source=chatgpt.com

29. Harris CR, Jarrod Millman K, van der Walt SJ, Gommers R, Virtanen P, Cournapeau D, et al. Array programming with NumPy. nature.comCR Harris, KJ Millman, SJ Van Der Walt, R Gommers, P Virtanen, D Cournapeau, E WieserNature, 2020•nature.com [Internet]. 2020 [cited 2025 Jul 9];585:357. Available from: https://www.nature.com/articles/s41586-020-2649-2

30. SciPy WM, 2010 undefined. Data structures for statistical computing in Python. pdfs.semanticscholar.orgW McKinneySciPy, 2010•pdfs.semanticscholar.org [Internet]. 2010 [cited 2025 Jul 9]; Available from: https://pdfs.semanticscholar.org/ef4e/f7f38bb907e5d7b4df3e6ff1db269d4970f5.pdf?_gl=1*1t91t2v*_ga*OTE5NDAxODkxLjE3MTcyMDIyOTM.*_ga_H7P4ZT52H5*MTcxNzIwMjI5My4xLjAuMTcxNzIwMjMwMy41MC4wLjA

31. Pedregosa FABIANPEDREGOSA F, Michel V, Grisel OLIVIERGRISEL O, Blondel M, Prettenhofer P, Weiss R, et al. Scikit-learn: Machine Learning in Python. Journal of Machine Learning Research [Internet]. 2011 [cited 2025 May 9];12(85):2825–30. Available from: http://jmlr.org/papers/v12/pedregosa11a.html

32. Chawla N V., Bowyer KW, Hall LO, Kegelmeyer WP. SMOTE: Synthetic Minority Over-sampling Technique. Journal of Artificial Intelligence Research [Internet]. 2002 Jun 1 [cited 2025 May 20];16:321–57. Available from: https://www.jair.org/index.php/jair/article/view/10302

33. McInnes L, Healy J, Melville J. UMAP: Uniform Manifold Approximation and Projection for Dimension Reduction. 2018 Feb 9 [cited 2025 Jul 7]; Available from: https://arxiv.org/pdf/1802.03426

34. Maaten L Van der, research GHJ of machine learning, 2008 undefined. Visualizing data using t-SNE. jmlr.org [Internet]. 2008 [cited 2025 Jul 7];9:2579–605. Available from: https://www.jmlr.org/papers/volume9/vandermaaten08a/vandermaaten08a.pdf?fbcl

35. Lundberg S, information SLA in neural, 2017 undefined. A unified approach to interpreting model predictions. proceedings.neurips.ccSM Lundberg, SI LeeAdvances in neural information processing systems, 2017•proceedings.neurips.cc [Internet]. [cited 2025 Jul 9]; Available from: https://proceedings.neurips.cc/paper/2017/hash/8a20a8621978632d76c43dfd28b67767-Abstract.html

36. Hunter JD. Matplotlib: A 2D graphics environment. Comput Sci Eng. 2007;9(3):90–5.

37. Software MWJ of OS, 2021 undefined. Seaborn: statistical data visualization. joss.theoj.orgML WaskomJournal of Open Source Software, 2021•joss.theoj.org [Internet]. [cited 2025 Jul 9]; Available from: https://joss.theoj.org/papers/10.21105/joss.03021.pdf

38. Flexible imputation of missing data. Second edition. | Stef van Buuren [Internet]. [cited 2025 May 7]. Available from: https://stefvanbuuren.name/publication/vanbuuren-2018/

39. Mogasale V, Maskery B, Ochiai RL, Lee JS, Mogasale V V., Ramani E, et al. Burden of typhoid fever in low-income and middle-income countries: A systematic, literature-based update with risk-factor adjustment. Lancet Glob Health [Internet]. 2014 Oct 1 [cited 2025 May 6];2(10):e570–80. Available from: https://pubmed.ncbi.nlm.nih.gov/25304633/

40. Background document: the diagnosis, treatment and prevention of typhoid fever [Internet]. [cited 2025 May 7]. Available from: https://iris.who.int/handle/10665/370492

41. Dyson ZA, Klemm EJ, Palmer S, Dougan G. Antibiotic Resistance and Typhoid. Clin Infect Dis [Internet]. 2019 Mar 7 [cited 2025 May 7];68(Suppl 2):S165. Available from: https://pmc.ncbi.nlm.nih.gov/articles/PMC6405283/

42. Laxminarayan R, Matsoso P, Pant S, Brower C, Røttingen JA, Klugman K, et al. Access to effective antimicrobials: A worldwide challenge. The Lancet [Internet]. 2016 Jan 9 [cited 2025 May 7];387(10014):168–75. Available from: https://pubmed.ncbi.nlm.nih.gov/26603918/

43. Crump JA, Luby SP, Mintz ED. The global burden of typhoid fever. Bull World Health Organ [Internet]. 2004 May [cited 2025 May 6];82(5):346. Available from: https://pmc.ncbi.nlm.nih.gov/articles/PMC2622843/

44. Hosoglu S, Geyik MF, Akalin S, Ayaz C, Kokoglu OF, Loeb M. A simple validated prediction rule to diagnose typhoid fever in Turkey. Trans R Soc Trop Med Hyg [Internet]. 2006 Nov [cited 2025 May 7];100(11):1068–74. Available from: https://pubmed.ncbi.nlm.nih.gov/16697432/

45. Biochemistry of Lipids, Lipoproteins and Membranes | ScienceDirect [Internet]. [cited 2025 May 8]. Available from: https://www.sciencedirect.com/book/9780444532190/biochemistry-of-lipids-lipoproteins-and-membranes

46. Ganz T. Anemia of Inflammation. New England Journal of Medicine [Internet]. 2019 Sep 19 [cited 2025 May 8];381(12):1148–57. Available from: https://pubmed.ncbi.nlm.nih.gov/31532961/

47. Semenza GL. Oxygen sensing, homeostasis, and disease. N Engl J Med [Internet]. 2011 Aug 11 [cited 2025 May 8];365(6):537–47. Available from: https://pubmed.ncbi.nlm.nih.gov/21830968/

48. Charles S, Edgar MP, Kasoma NA. The Hunt for Antipox Compounds against Monkeypox Virus Thymidylate Kinase and Scaffolding Protein Leveraging Pharmacophore Modeling, Molecular Docking, ADMET Studies and Molecular Dynamics Simulation Studies. Virology & Mycology [Internet]. 2023 Nov 2 [cited 2024 Jan 12];12(4):1–14. Available from: https://www.longdom.org/open-access/the-hunt-for-antipox-compounds-against-monkeypox-virus-thymidylate-kinase-and-scaffolding-protein-leveraging-pharmacophore-modelin-104682.html

49. David CC, Jacobs DJ. Principal component analysis: A method for determining the essential dynamics of proteins. Methods in Molecular Biology. 2014;1084:193–226.

50. Wold S, Esbensen K, Geladi P. Principal component analysis. Chemometrics and Intelligent Laboratory Systems [Internet]. 1987 Aug 1 [cited 2025 May 9];2(1–3):37–52. Available from: https://www.sciencedirect.com/science/article/abs/pii/0169743987800849?via%3Dihub

51. Almadhoun MB, Burhanuddin M. Optimizing Feature Selection and Machine Learning Algorithms for Early Detection of Prediabetes Risk: Comparative Study. JMIR Bioinform Biotech [Internet]. 2025 Jul 31 [cited 2025 Aug 26];6:e70621. Available from: https://pmc.ncbi.nlm.nih.gov/articles/PMC12314567/

52. Varma S, Simon R. Bias in error estimation when using cross-validation for model selection. BMC Bioinformatics 2006 7:1 [Internet]. 2006 Feb 23 [cited 2025 Nov 29];7(1):91-. Available from: https://link.springer.com/article/10.1186/1471-2105-7-91

53. Bergstra J, Ca JB, Ca YB. Random search for hyper-parameter optimization. The Journal of Machine Learning Research [Internet]. 2012 Feb 1 [cited 2025 Nov 29];13:281–305. Available from: https://dl.acm.org/doi/pdf/10.5555/2188385.2188395

54. Nakagawa S, Johnson PCD, Schielzeth H. The coefficient of determination R2 and intra-class correlation coefficient from generalized linear mixed-effects models revisited and expanded. J R Soc Interface [Internet]. 2017 Sep 1 [cited 2025 Nov 29];14(134). Available from: 10.1098/rsif.2017.0213

55. Krstajic D, Buturovic LJ, Leahy DE, Thomas S. Cross-validation pitfalls when selecting and assessing regression and classification models. Journal of Cheminformatics 2014 6:1 [Internet]. 2014 Mar 29 [cited 2025 Nov 29];6(1):10-. Available from: https://link.springer.com/article/10.1186/1758-2946-6-10

56. Tibshirani RJ, Tibshirani R. A bias correction for the minimum error rate in cross-validation. 10.1214/08-AOAS224 [Internet]. 2009 Jun 1 [cited 2025 Nov 29];3(2):822–9. Available from: https://projecteuclid.org/journals/annals-of-applied-statistics/volume-3/issue-2/A-bias-correction-for-the-minimum-error-rate-in-cross/10.1214/08-AOAS224.full

57. Parry CM, Vinh H, Chinh NT, Wain J, Campbell JI, Hien TT, et al. The Influence of reduced susceptibility to fluoroquinolones in Salmonella enterica serovar Typhi on the clinical response to ofloxacin therapy. PLoS Negl Trop Dis. 2011;5(6).

58. Marchello C, Birkhold M, Infection JCJ of, 2020 undefined. Complications and mortality of typhoid fever: a global systematic review and meta-analysis. Elsevier [Internet]. [cited 2025 Jul 7]; Available from: https://www.sciencedirect.com/science/article/pii/S0163445320306903

59. Wain J, Song Diep T, Anh VH, Walsh AM, Thi Tuyet Hoa N, Parry CM, et al. Quantitation of Bacteria in Blood of Typhoid Fever Patients and Relationship between Counts and Clinical Features, Transmissibility, and Antibiotic Resistance [Internet]. Vol. 36, JOURNAL OF CLINICAL MICROBIOLOGY. 1998. Available from: https://journals.asm.org/journal/jcm

60. Levine M, Black R, Ferreccio C, Lancet RGT, 1987 undefined. Large-scale field trial of Ty21a live oral typhoid vaccine in enteric-coated capsule formulation. ElsevierMM Levine, RE Black, C Ferreccio, R Germanier, CT CommitteeThe Lancet, 1987•Elsevier [Internet]. [cited 2025 Jul 7]; Available from: https://www.sciencedirect.com/science/article/pii/S0140673687904806

61. Charles S, Pius Edgar M, Mahapatra RK. Host-Directed Anti-Fusion Aptamers and Small Molecules as Respiratory Syncytial Virus (RSV) Inhibitors: An in silico-based study. Journal of Biotechnology and Biomedicine. 2024;7:485–97.

62. Xu Y, Goodacre R. On Splitting Training and Validation Set: A Comparative Study of Cross-Validation, Bootstrap and Systematic Sampling for Estimating the Generalization Performance of Supervised Learning. J Anal Test [Internet]. 2018 Jul 1 [cited 2025 Jun 30];2(3):249–62. Available from: https://pubmed.ncbi.nlm.nih.gov/30842888/

62. Efron, B., & Tibshirani, R. J. (1993). An Introductio… – Google Scholar [Internet]. [cited 2025 Jul 7]. Available from: https://scholar.google.com/scholar?hl=en&as_sdt=0%2C5&q=%E2%80%A2%09Efron%2C+B.%2C+%26+Tibshirani%2C+R.+J.+%281993%29.+An+Introduction+to+the+Bootstrap.+Chapman+%26+Hall%2FCRC.&btnG=

64. Abbas S, Sattar A, Shah SH, Hafeez S, Mahmood W, Iqbal R, et al. THE ROLE OF ARTIFICIAL INTELLIGENCE IN PERSONALIZED MEDICINE AND PREDICTIVE DIAGNOSTICS–A NARRATIVE REVIEW. insightsjhr.com [Internet]. 2025 [cited 2025 Jul 7]; Available from: https://insightsjhr.com/index.php/home/article/view/510

65. Antillón M, Warren JL, Crawford FW, Weinberger DM, Kürüm E, Pak GD, et al. The burden of typhoid fever in low-and middle-income countries: a meta-regression approach. journals.plos.orgM Antillón, JL Warren, FW Crawford, DM Weinberger, E Kürüm, GD Pak, F Marks, VE PitzerPLoS neglected tropical diseases, 2017•journals.plos.org [Internet]. 2017 Feb 27 [cited 2025 Jul 7];11(2). Available from: https://journals.plos.org/plosntds/article?id=10.1371/journal.pntd.0005376

66. Van Calster B, McLernon DJ, Van Smeden M, Wynants L, Steyerberg EW, Bossuyt P, et al. Calibration: The Achilles heel of predictive analytics. BMC Med [Internet]. 2019 Dec 16 [cited 2025 Jul 8];17(1):1–7. Available from: https://bmcmedicine.biomedcentral.com/articles/10.1186/s12916-019-1466-7

67. Niculescu-Mizil A, … RC of the 22nd international conference on, 2005 undefined. Predicting good probabilities with supervised learning. dl.acm.org [Internet]. 2005 [cited 2025 Jul 8];625–32. Available from: 10.1145/1102351.1102430

68. review WGM weather, 1950 undefined. Verification of forecasts expressed in terms of probability. books.google.comWB GlennMonthly weather review, 1950•books.google.com [Internet]. [cited 2025 Jul 12]; Available from: https://books.google.com/books?hl=en&lr=&id=jnbpAAAAMAAJ&oi=fnd&pg=RA1-PA1&dq=%E2%80%A2%09Brier,+G.+W.+(1950).+Verification+of+forecasts+expressed+in+terms+of+probability.+Monthly+Weather+Review,+78(1),+1%E2%80%933.+ 10.1175/1520-0493(1950)078%3C0001:VOFEIT%3E2.0.CO%3B2&ots=0Y1ZZHjIvR&sig=Nwq2If5MxzoY8cZjfYgkY86UnRg

69. Austin P, medical ESS methods in, 2017 undefined. Events per variable (EPV) and the relative performance of different strategies for estimating the out-of-sample validity of logistic regression models. journals.sagepub.comPC Austin, EW SteyerbergStatistical methods in medical research, 2017•journals.sagepub.com [Internet]. 2017 Apr 1 [cited 2025 Jul 12];26(2):796–808. Available from: https://journals.sagepub.com/doi/abs/10.1177/0962280214558972

70. Lundberg S, information SLA in neural, 2017 undefined. A unified approach to interpreting model predictions. proceedings.neurips.ccSM Lundberg, SI LeeAdvances in neural information processing systems, 2017•proceedings.neurips.cc [Internet]. [cited 2025 Jul 7]; Available from: https://proceedings.neurips.cc/paper/2017/hash/8a20a8621978632d76c43dfd28b67767-Abstract.html

71. Molnar C. Interpretable machine learning [Internet]. 2020 [cited 2025 Jul 7]. Available from: https://books.google.com/books?hl=en&lr=&id=jBm3DwAAQBAJ&oi=fnd&pg=PP1&dq=3.%09Molnar,+C.+(2022).+interpretable+Machine+Learning:+A+Guide+for+Making+Black+Box+Models+Explainable+(2nd+ed.).+ https://christophm.github.io/interpretable-ml-book/&ots=EhwRUiFKY5&sig=HgXrqnPQjLQtOWN-3zq6lRcfsc0

72. Stanaway JD, Reiner RC, Blacker BF, Goldberg EM, Khalil IA, Troeger CE, et al. The global burden of typhoid and paratyphoid fevers: a systematic analysis for the Global Burden of Disease Study 2017. Lancet Infect Dis. 2019 Apr 1;19(4):369–81.

73. Tamelytė E, Vaičekauskienė G, Dagys A, Lapinskas T, Jankauskaitė L. Early Blood Biomarkers to Improve Sepsis/Bacteremia Diagnostics in Pediatric Emergency Settings. Medicina 2019, Vol 55, Page 99 [Internet]. 2019 Apr 10 [cited 2025 Sep 5];55(4):99. Available from: https://www.mdpi.com/1648-9144/55/4/99/htm

74. Ahmad M, Eckert C, ACM ATP of the 2018, 2018 undefined. Interpretable machine learning in healthcare. dl.acm.org [Internet]. 2018 Aug 15 [cited 2025 Jul 8];559–60. Available from: https://dl.acm.org/doi/abs/10.1145/3233547.3233667

74. Kawamoto K, Houlihan CA, Balas EA, Lobach DF. Improving clinical practice using clinical decision support systems: a systematic review of trials to identify features critical to success. BMJ [Internet]. 2005 Mar 31 [cited 2025 Jul 12];330(7494):765. Available from: https://www.bmj.com/content/330/7494/765

76. Shortliffe EH, Cimino JJ. Biomedical informatics: Computer applications in health care and biomedicine: Fourth edition. Biomedical Informatics: Computer Applications in Health Care and Biomedicine: Fourth Edition. 2014 Jan 1;1–965.

77. Rajkomar A, Dean J, Kohane I. Machine Learning in Medicine. New England Journal of Medicine [Internet]. 2019 Apr 4 [cited 2025 Jun 30];380(14):1347–58. Available from: https://pubmed.ncbi.nlm.nih.gov/30943338/

78. Topol, E. (2019). Deep Medicine: How Artificial Intellige… – Google Scholar [Internet]. [cited 2025 Jul 12]. Available from: https://scholar.google.com/scholar?hl=en&as_sdt=0%2C5&q=Topol%2C+E.+%282019%29.+Deep+Medicine%3A+How+Artificial+Intelligence+Can+Make+Healthcare+Human+Again.+Basic+Books.&btnG=

79. Jiang H, Kim Google Brain B, Guan MY, Gupta Google Research M. To trust or not to trust a classifier. proceedings.neurips.ccH Jiang, B Kim, M Guan, M GuptaAdvances in neural information processing systems, 2018•proceedings.neurips.cc [Internet]. [cited 2025 Jul 12]; Available from: https://proceedings.neurips.cc/paper/2018/hash/7180cffd6a8e829dacfc2a31b3f72ece-Abstract.html

80. Charles S, Edgar MP. Geometric Deep learning Prioritization and Validation of Cannabis Phytochemicals as Anti-HCV Non-nucleoside Direct-acting Inhibitors. Biomed Eng Comput Biol [Internet]. 2024 Jan 12 [cited 2025 Apr 23];15. Available from: https://journals.sagepub.com/doi/10.1177/11795972241306881

81. Charles S, Edgar MP, Mahapatra RK. Artificial intelligence based virtual screening study for competitive and allosteric inhibitors of the SARS-CoV-2 main protease. J Biomol Struct Dyn. 2023;

82. Dellit TH, Owens RC, Mcgowan JE, Gerding DN, Weinstein RA, Burke JP, et al. Infectious Diseases Society of America and the Society for Healthcare Epidemiology of America guidelines for developing an institutional program to enhance. academic.oup.comTH Dellit, RC Owens, JE McGowan, DN Gerding, RA Weinstein, JP Burke, WC HuskinsClinical infectious diseases, 2007•academic.oup.com [Internet]. [cited 2025 Jul 13]; Available from: https://academic.oup.com/cid/article-abstract/44/2/159/328413

83. Spellberg B, Guidos R, Gilbert D, Bradley J, Boucher HW, Scheld WM, et al. The epidemic of antibiotic-resistant infections: a call to action for the medical community from the Infectious Diseases Society of America. academic.oup.comB Spellberg, R Guidos, D Gilbert, J Bradley, HW Boucher, WM Scheld, JG BartlettClinical infectious diseases, 2008•academic.oup.com [Internet]. [cited 2025 Jul 13]; Available from: https://academic.oup.com/cid/article-abstract/46/2/155/452327

84. Wong V, Baker S, Pickard D, Parkhill J, … APN, 2015 undefined. Phylogeographical analysis of the dominant multidrug-resistant H58 clade of Salmonella Typhi identifies inter– and intracontinental transmission events. nature.comVK Wong, S Baker, DJ Pickard, J Parkhill, AJ Page, NA Feasey, RA Kingsley, NR ThomsonNature genetics, 2015•nature.com [Internet]. [cited 2025 Jul 13]; Available from: https://www.nature.com/articles/ng.3281

85. Niculescu-Mizil A, Caruana R. Predicting good probabilities with supervised learning. ICML 2005 – Proceedings of the 22nd International Conference on Machine Learning [Internet]. 2005 [cited 2025 Oct 19];625–32. Available from: 10.1145/1102351.1102430?download=true

86. Ojeda FM, Jansen ML, Thiéry A, Blankenberg S, Weimar C, Schmid M, et al. Calibrating machine learning approaches for probability estimation: A comprehensive comparison. Stat Med [Internet]. 2023 Dec 20 [cited 2025 Oct 19];42(29):5451–78. Available from: 10.1002/sim.9921

87. Tyralis H, Papacharalampous G. A review of predictive uncertainty estimation with machine learning. Artif Intell Rev [Internet]. 2024 Apr 1 [cited 2025 Oct 19];57(4):1–65. Available from: https://link.springer.com/article/10.1007/s10462-023-10698-8

